# Ethical and Social Considerations of Applying Artificial Intelligence in Healthcare; a Two-Pronged Scoping Review

**DOI:** 10.1101/2025.02.10.25321994

**Authors:** Emanuele Ratti

**Affiliations:** University of Bristol Michael Morrison, University of Oxford Ivett Jakab, YAGHMA B.V

**Author notes:** Clinical trial number: not applicable.

**Keywords:** artificial intelligence, medicine, healthcare, scoping review

## Abstract

**Background:** Artificial Intelligence (AI) is being designed, tested, and in many cases actively employed in almost every aspect of healthcare from primary care to public health. It is by now well established that any application of AI carries an attendant responsibility to consider the ethical and societal aspects of its development, deployment and impact. However, in the rapidly developing field of AI, developments such as machine learning, neural networks, generative AI, and large language models have the potential to raise new and distinct ethical and social issues compared to, for example, automated data processing or more ‘basic’ algorithms.

**Methods:** This article presents a scoping review of the ethical and social issues pertaining to AI in healthcare, with a novel two-pronged design. One strand of the review (SR1) consists of a broad review of the academic literature restricted to a recent timeframe (2021-23), to better capture up to date developments and debates. The second strand (SR2) consists of a narrow review, limited to prior systematic and scoping reviews on the ethics of AI in healthcare, but extended over a longer timeframe (2014-2024) to capture longstanding and recurring themes and issues in the debate. This strategy provides a practical way to deal with an increasingly voluminous literature on the ethics of AI in healthcare in a way that accounts for both the depth and evolution of the literature.

**Results:** SR1 captures the heterogeneity of audience, medical fields, and ethical and societal themes (and their tradeoffs) raised by AI systems. SR2 provides a comprehensive picture of the way scoping reviews on ethical and societal issues in AI in healthcare have been conceptualized, as well as the trends and gaps identified.

**Conclusion:** Our analysis shows that the typical approach to ethical issues in AI, which is based on the appeal to general principles, becomes increasingly unlikely to do justice to the nuances and specificities of the ethical and societal issues raised by AI in healthcare, as the technology moves from abstract debate and discussion to real world situated applications and concerns in healthcare settings.

## 1 BACKGROUND

### 1.1 Context

Artificial intelligence (AI) systems have the potential for wide-ranging and transformative impact on healthcare, including both primary and secondary care settings (Panch et al 2019; Topol 2019). AI systems, which we define broadly to include ‘basic’ algorithms as well as more advanced digital computational techniques such as machine learning, neural networks, and Large Language Models et cetera, offer faster, more accurate and effective diagnosis, prognosis, risk assessment, and triage decisions, enable precision medicine, remote monitoring, patient support, and more. However, the risks of AI are also well documented; the potential for discriminatory, unfair or socially detrimental outcomes, lack of accountability and appropriate oversight, erosion of professional expertise, threats to privacy and confidentiality, and overhyped claims (Fenech et al 2018; Kaltheuner 2021; Aquino et al 2023; Fazelpour and Danks 2021). As a result, there has been a concomitant proliferation of works on the ethical and societal aspects of artificial intelligence in healthcare^2^ (Cartolovini et al 2022; Goirand et al 2021; Kariman, Petelos and Evers 2022; Mollmann, Mirbabaie and Stieglitz 2021; Morley et al 2020; Murphy et al 2021; Naik et al 2022).^3^

Technical and scientific reports on the real and potential application of AI in healthcare are both long standing, dating back to at least the mid 1990s and have been subject to rapid growth and acceleration in the volume of material published in recent years (see Guo et al 2020 and Qin et al 2022 for scientometric analyses of the field). In a similar fashion, published discussion on the ethics of AI in healthcare has developed from general issues of concern (Tsamados et al 2021) to more in-depth explorations of ethical and societal issues attending application of AI in specific subfields of medicine such as rare disease (Hallowell et al 2023) or mental health (Skorburg, O’Doherty, & Friesen 2024), and particular types of algorithmic agent such as Clinical Decision Support Systems and chatbots (Braun et al 2021; Fournier-Tombs & McHardy, 2023).

In order to understand what ethical and societal issues are at stake in this fast moving world of AI in healthcare, ideally one could conduct a traditional systematic review. This would offer a fully comprehensive account of the literature on ethics of AI in healthcare. However, due to the size of the published literature such an approach is impractical to undertake in terms of resources and time. For example Benefo et al (2023) identified 1028 publications on ethical, legal and social implications of AI in healthcare published 1991-2020 from the Clarivate ISI Web of Science database alone. Fortunately, there are types of systematic reviews that are tailored to the goal of providing a ‘higher-level’ view of fast changing fields like AI ethics. In particular, scoping reviews are excellent for this task.

Scoping reviews, as defined by the Canadian Institute of Health Research, are reviews “that systematically map the literature available on a topic, identifying key concepts, theories, sources of evidence and gaps in research”^4^, but unlike traditional systematic reviews (such as meta-analyses), they are more exploratory in nature (Peters et al 2020). The suitability of the scoping review approach to the ethics of AI in healthcare is reinforced by the fact that a number of similar reviews have already been published (Cartolovini et al 2022; Goirand et al 2021; Morley et al 2020; Murphy et al 2021). However, the rapidly evolving nature of AI in healthcare and the transition from general concerns to more detailed analyses of the ethical and societal issues in specific medical subfields or types of application, means that new scoping reviews are needed, and indeed will continue to be needed in future, to keep abreast of this ever changing landscape. Moreover, the rise of novel computational systems such as Large Language Models and generative AI means that the ethical and societal issues raised by AI are not static but are also evolving. Here we propose a scoping review of the issues of ethical and societal relevance raised by AI in healthcare, with a novel design intended to account for the longstanding but rapidly growing and diversifying body of literature in this field. The present scoping review comprises two parallel strands of activity. The first strand consists of a ‘broad’ scoping review of ethical and societal issues raised by AI in the medical context (SR1). In order to return a manageable amount of data this review is limited in scope to recent years. The second strand entails a ‘deep’ scoping review, which extends over a greater time period, but it is limited to existing scoping reviews with a published methodology^5^ (SR2). This novel structure was motivated by the fact that there have been several recent scoping reviews on ethical and societal issues raised by AI in healthcare. For this reason, our ‘broad’ review will cover only the most recent years, since the previous years have been already covered by existing reviews. At the same time, we also want to understand how ethical and societal issues mapped by existing scoping reviews have been conceptualised. In this way our research strategy aims to account for both the historical depth of scholarship on ethics of AI in healthcare and the cutting edge of contemporary ethical debates on the topic.

### 1.2 Objectives

Both strands of the review (SR1 and SR2) share the same objectives, which are; (1) to examine the extent and nature of research on ethical and societal issues raised by AI in the medical context; (2) to organise this research by well-specified themes; and (3) to identify important gaps in this research. In addition to characterising the discussions on ethical and societal issues raised by AI in medicine, our analysis shows that these issues are becoming more and more specialised, and that a principle-based approach that has characterised AI ethics so far (Floridi and Cowls 2019) is uninformative, and not useful to practitioners who are struggling to design and implement AI tools in the medical context.

## 2. METHODS

Our approach to scoping reviews is informed by agreed methodological guidelines, pioneered by (Arksey and O’Malley 2005) and standardised in Peters et al (2020). For both SR1 and SR2, we have followed the five-stage procedures described in Arksey and O’Malley (2005), consisting of:

1. establishing the research question
2. identifying relevant studies
3. selecting the studies
4. charting data
5. reporting collated results

### 2.1 The research question

Our interest lies in mapping the debate on the ethical and societal issues raised by AI technologies in the healthcare context. We decided not to limit our analysis to specific AI tools like machine learning or large language models or to one domain of medicine (e.g. by including clinical care but not public health), but designed our search strings to encompass a broad range of computational technologies across all areas of health and care. This is also reflected in the keywords used (see 2.2 below). More precisely, our research question is as follows:

#### What are the ethical and societal issues raised by computational technologies in medicine that are discussed in the literature?

In order to distinguish between ‘ethical’ and ‘societal’ issues, we utilise the scheme offered by (Ferdman and Ratti 2024), where a distinction is drawn between questions pertaining the impact of technologies on individuals, and questions pertaining the impact of technologies at the level of populations or groups including groups of experts such as medical professionals, policy makers, or engineers. The former set of questions are said to be ‘ethical’, while the latter are considered to be ‘societal’. INote that this approach is not meant to represent any type of fundamental distinction; it is simply a useful heuristic to allow us to deal practically with the ethics and politics of technology. It is important to notice that sometimes ethical and societal issues raised by AI systems are entangled with their epistemic dimensions, as in the case of explainability and accuracy. This is especially because of how epistemic considerations shape the ethical and societal effects of AI systems, as we will show.

The research question is the same in both SR1 and SR2, even though the focus might be considered slightly different, as we will show when discussing inclusion and exclusion criteria. In the case of SR1, the ‘broad review’, the ‘literature’ to be returned in response to the research question is provided by recent peer reviewed research articles and commentaries, while in the case of SR2, the ‘deep review’ the ‘literature’ is limited to prior scoping reviews but published over a longer timeframe.

### 2.2 Search strategy and inclusion/exclusion criteria (study identification)

The identification of relevant studies was done by selecting the type of literature to include; the databases used to look for the literature; and the search strings developed and employed to identify relevant studies in the selected databases. SR1 is intended to capture a broad scope of recent research. Accordingly, it is limited to peer-reviewed articles published between 2021 and 2023. SR2 is the ‘narrow’ but deep review and is limited to scoping and systematic reviews published between 2014 and 2024. We excluded other literature reviews such as narrative or unstructured reviews because they are difficult to summarise and extract data from. We have excluded grey literature, book reviews, book chapters, books, codes of conduct, and policy documents because the extant literature is simply too large to be manageable in a reasonable timeframe.

The literature searches for SR1 and SR1 were conducted on the following online electronic databases. For both searches, Ovid Medline was selected to address the medical context (which is central to our research question), while IEEE was chosen to address the engineering and computer science domain pertinent to AI. In addition, ISI Web of Science, was used to provide broad coverage while also capturing relevant social science literature on AI in healthcare. For SR1 we also consulted Philpapers, a database specifically focused on philosophy journals. This is because of the relevance of specific philosophy journals in the debate on the ethical and societal issues in medical AI. We have not considered Philpapers for SR2, given that scoping reviews are typically not published in philosophy journals.

Keywords for SR1 search strings were structured into three levels: the technology used, the context, and the focus. This is motivated by the fact that our research question is about the use of AI (i.e. a technology) in medicine (the context) and the ethical and societal issues it raises (the focus). For each level, various options separated by the ‘OR ’operator were included (see Appendix 1). Each level was connected to the others by the operator ‘AND’. Additional restrictions were added for specific databases based on the volume of results returned and the options offered by the search interface of each database; for example on Web of Science, the research was limited to journal articles only and to English language publications only. Searches were run to account for years 2021, 2022, and 2023 (up to and including the date on which each search was run). Specific SR1 search strings for each database are shown in Table s1 of Appendix 1.

Keywords for SR2 were organised in a similar way as SR1, with two main differences: The timeframe of the review was extended to encompass 2014 to 2024, in accordance with the idea of a ‘deep review’ spanning a longer time period to capture the history of the debate, and the scope of the review was limited to results with the keywords ‘systematic’ or ‘scoping’ and ‘review’ in the title or abstract since the ‘deep review’ was to focus exclusively on prior review articles. This was done because existing reviews were identified as an efficient way of capturing the large historic volume of literature on the ethics of AI in healthcare whilst generating a manageable data set. Table s2 in Appendix 1 contains the search strings and results for SR2.

### 2.3 Study Selection

Studies for SR1 were screened in the following way. After the preliminary search described in 2.2, results from each database were inserted into a reference management system (EndNote), and moved to Rayyan, an online platform designed to facilitate double blind screening of research articles (Ouzzni et al 2016). Once all results from the four databases searched were uploaded to Rayyan, the platform’s internal ‘duplicate screening’ tool was used to remove duplicate references. This tool is partially automated, but allows users to check each set of duplicates before a final decision is made, adding a greater level of control about what is or is not removed. Following this step, the remaining publications in SR1 were independently screened by ER and MM by looking at the title and abstract only^6^. Publications were excluded according to the following exclusion criteria:

- any remaining undetected duplicate papers.
- articles where ethical and societal issues in medical AI were not the primary focus (including primarily technical accounts of AI development or testing in healthcare setting that did not discuss ethical or social issues or mentioned them only superficially)
- articles seemingly about either AI in general (with no mention of medicine) or about medicine (with no reference to AI).
- systematic reviews
- non-English language publications
- Articles dealing mainly with medical robotics or wearable devices, even if the ethical and social issues are addressed.

Medical robotics and wearable devices were excluded, partly as a pragmatic decision to keep the volume of papers returned manageable, but also because these topics, whilst they do involve AI, foreground additional issues that are beyond the scope of this review. The ethical and societal issues raised by medical robots implicate the physical presence of a robot and its impact on robot-patient interactions (Datteri 2013; Sharkey & Sharkey 2012), which the digital algorithms that concern us here, largely do not. Data from wearable devices, such as smartwatches and fitness trackers, is certainly subject to algorithmic processing, but wearables themselves are again physical devices primarily concerned with generating data. They raise additional concerns around surveillance, intrusiveness, and reliability of ‘self-generated’ healthcare information (Capulli et al 2025), which again are less pertinent to this review.

After this screening, MM and ER agreed on a list of 193 publications. In charting data, which involved a full text reading of the papers, a further round of exclusions was conducted as for example some papers had an English language abstract but the main text was in another language, or an abstract that mentioned ethical or societal issues turned out to belong to a largely technical paper with a small section on e.g. receiving ethical approval for a study. This final round of exclusions gives the final number of included studies in SR1 as 164 (see Figure 1).

**Figure 1.**
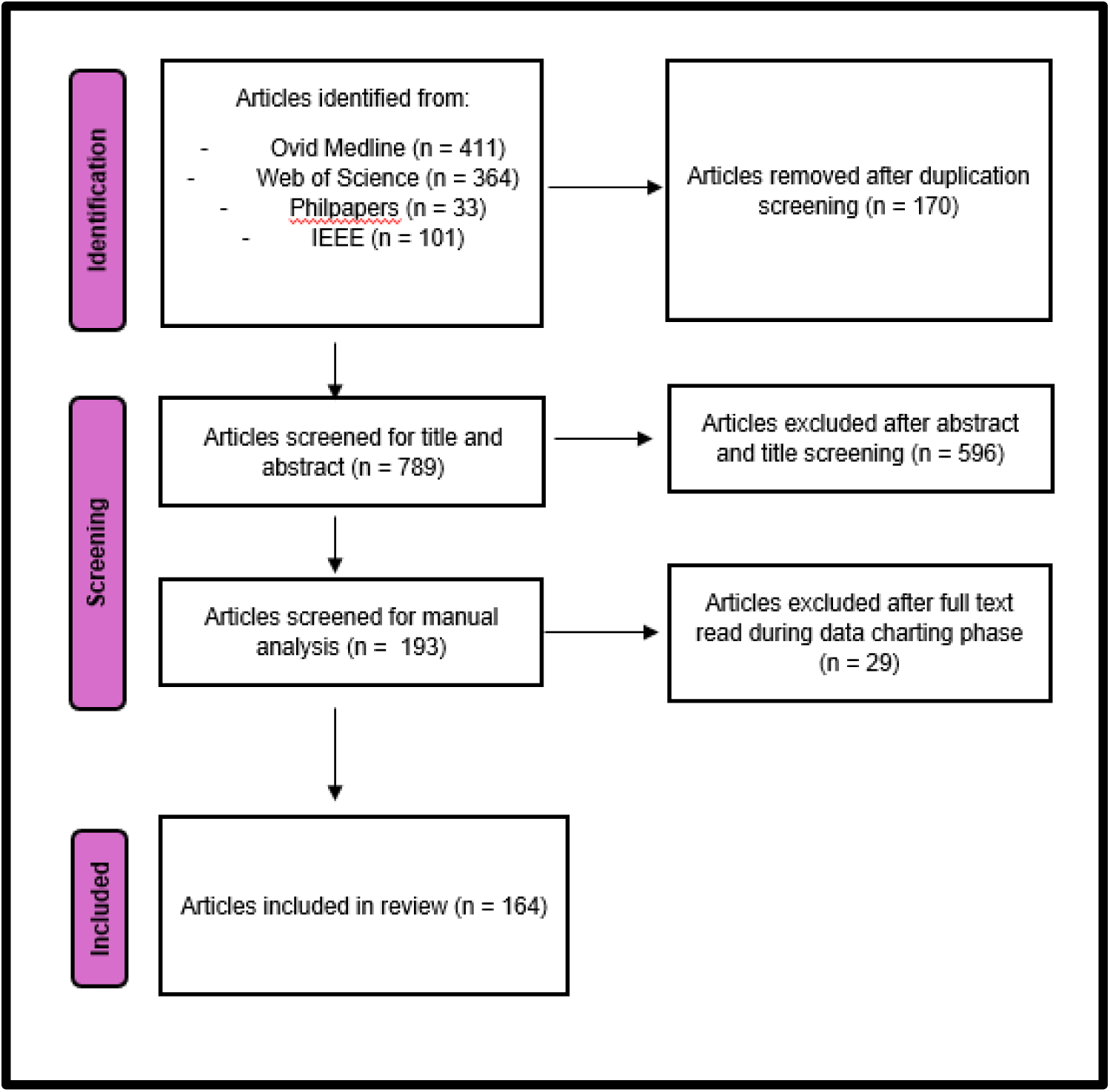
PRISMA 2020 Flow Diagram for SR1

The screening of studies for SR2 followed a similar strategy. Results from each database were again moved to EndNote and collectively uploaded to Rayyan, where the detect duplicates tool was applied. The remaining results were then independently screened by ER, MM, and IJ by looking at titles and abstracts only. Conflicting decisions between reviewers were resolved iteratively through a series of meetings to build consensus on defining and applying our criteria. Publications were excluded according to the following criteria:

- any remaining undetected duplicate papers
- reviews seemingly not related to ethical and societal issues in medical AI
- reviews seemingly about either AI in general (with no mention of medicine) or about medicine (with no reference to AI)
- Articles dealing mainly with medical robotics or wearable devices, even if the ethical and social issues are addressed.
- non-English language publications
- non-systematic or scoping reviews
- Articles with no clear or well-described published methodology

After this screening, ER, MM, and IJ agreed on a final list of 23 publications. In charting the data, three additional publications were excluded, again based on examination of the full text (see Figure 2).

**Figure 2.**
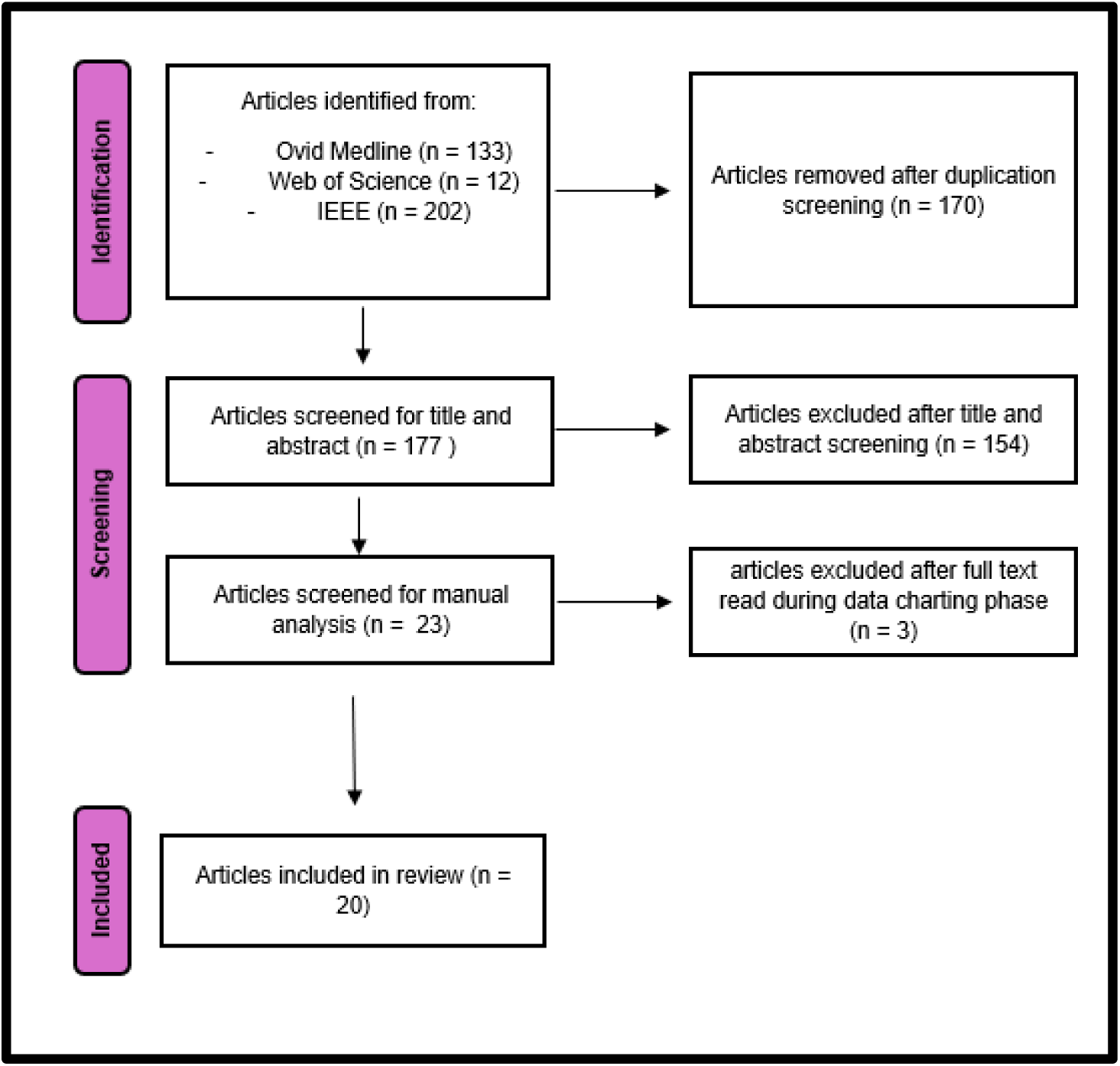
PRISMA 2020 Flow Diagram for SR2

### 2.4 Data charting

For each of SR1 and SR2 a separate chart was designed to collect and organise extracted data and metadata from each of the included papers for that strand of the review. The tables were developed iteratively and reflexively. For SR1, ER and MM independently extracted data from 20 studies using a preliminary table, and determined whether the data added to the categories in the preliminary chart were consistent with answering the research question. Following a collective review of this text charting, ER, MM, and IJ refined the list of variables to extract over several further iterations before agreeing on the final format for Chart 1, as presented in Appendix 2. Chart 1 collects descriptive metadata on each paper in the SR1 dataset, in the form of publication year, authors, title, journal, and research question-relevant information. This latter category includes a number of important descriptive and conceptual components of the literature analysed. First, articles about the ethical and societal issues raised by the use of AI in medicine were classified as either non-empirical (e.g. commentaries, conceptual analyses, etc) or empirical (i.e. studies using qualitative methodologies such as semi-structured interviews). Where articles were categorised as empirical, additional descriptive characteristics such as study population, sample size, empirical methodology and so on, were taken into account. Second, articles were categorised as either considering medicine in a general or broad sense, or as considering ethical or societal aspects of AI in a specific domain of healthcare such as surgery, cardiology, public health et cetera. These domains were derived from a preliminary list, with an ‘other’ category allowing us to record unanticipated or uncommon subdomains. Similarly, the technological focus of each paper was addressed, categorising papers by area of (current or potential) application using the following four categories: ‘diagnostic and treatment applications’, ‘patient engagement and adherence applications’, ‘administrative application’s (e.g. scheduling of medical visits or procedures, automated billing), or ‘other’ to capture miscellaneous or ambiguous areas of application.

Given that identifying and mapping the ethical and societal issues raised by the use of AI in healthcare was the major focus of the research question, a significant amount of effort went into extracting this data from each paper and organising it. This was achieved by all three authors identifying a set of key themes, based on *a priori* knowledge of major ethical principles and supplemented by exploratory reading of the literature on the ethics of AI. Each theme captures one issue or dimension within the broader normative debates on AI in healthcare. The selected themes, and the meanings of each of these terms and the way they were interpreted and applied by the authors in the course of the analysis is shown in Appendix 3. It was clear from the outset that we would need to anticipate and accommodate unexpected or novel ethical and social issues that arose during the course of charting the data. To this end an additional column of ‘other’ issues was created, acting as a ‘catch all’ category for anything that did not fit in one or more of our predefined categories. At the end of the analysis, the material captured in the ‘other’ column was then revisited, discussed and further categorised by all three authors (see section 4 and Appendix 6)..

A further categorization step involved assessing the intended audience for each paper; for example, was the paper aimed primarily at healthcare professionals, or to other audiences such as patients, developers, or regulators. To reduce subjectivity of decision making and inter-coder variability, only papers that clearly stated a target audience were classified in this way. For those articles that did not have a clearly identifiable or stated audience a ‘general’ category was also included as an option. Lastly, we were also attentive to whether the articles discussed the possibility of conflicts between specific values in discussing ethical and societal challenges of implementing AI in medicine (e.g. whether transparency conflicts with privacy).

SR2 followed a similar process: ER, MM, and IJ independently extracted data from the 23 selected publications (and excluded 3 as a result of this process, as shown in 2.3). Again, using an iterative process of chart development and review, a second data chart, Chart 2 was established (see Appendix 4). Chart 2 collected the same descriptive metadata on each paper (authors, title, journal, publication year), and a further set of descriptive metadata about each review, pertaining to the type of methodology used, the databases searched, whether grey literature is included, the search timeframe and duration (no of years covered), the area of medicine, the type of AI considered, and the research question addressed by the review^7^. Next, we reported the ethical and societal values that were identified by each scoping review as being discussed in the literature, and the trends and the trade-offs observed by the authors of the scoping review. Unlike SR1, for the deep review we did not specify a list of ethical themes in advance. Instead, we used an approach that is more data-driven, and reported the exact themes that the scoping reviews discussed using the terminology presented in that particular paper. Importantly, previous scoping reviews often identify gaps in the literature, and we think that the gaps identified provide a good indication of which ethical issues the authors think are important, so these were also recorded in Chart 2 (Appendix 4).

## 3 RESULTS

### 3.1 Timeframe

SR1 is focused on articles published between 2021 (n = 68), 2022 (n = 61), and 2023 (n = 34) (Figure 3). The discrepancy between 2023 and the other years is due to data for 2021 and 2022 comprising the full year while that for 2023 contains only articles published until the searches were conducted (August 2023). For SR2, we found articles as early as 2018 (n = 2), while most scoping reviews were published in 2022 (n = 7) and 2023 (n = 5). The timeframes covered by the scoping and systematic literature reviews in SR2 also show a wide variety in duration and date ranges, with the earliest search commencing in 1946 and the most recent concluding in 2023 (Figure 4). The number of articles involved also returned significantly different volumes of papers for each scoping review, ranging from 8 as the smallest data set to 263 papers in the largest. This is because each scoping review included in SR2 addressed a different research question and mobilised different keywords and search parameters. The full list of date range, research question, and volume of results for scoping reviews returned in SR2 are presented in Appendix 5. Please note that the full list of included papers in SR1 is not included in this paper, considering the high number of included articles and standards of reporting for such extensive findings. However, the list is available upon request.

**Figure 3.**
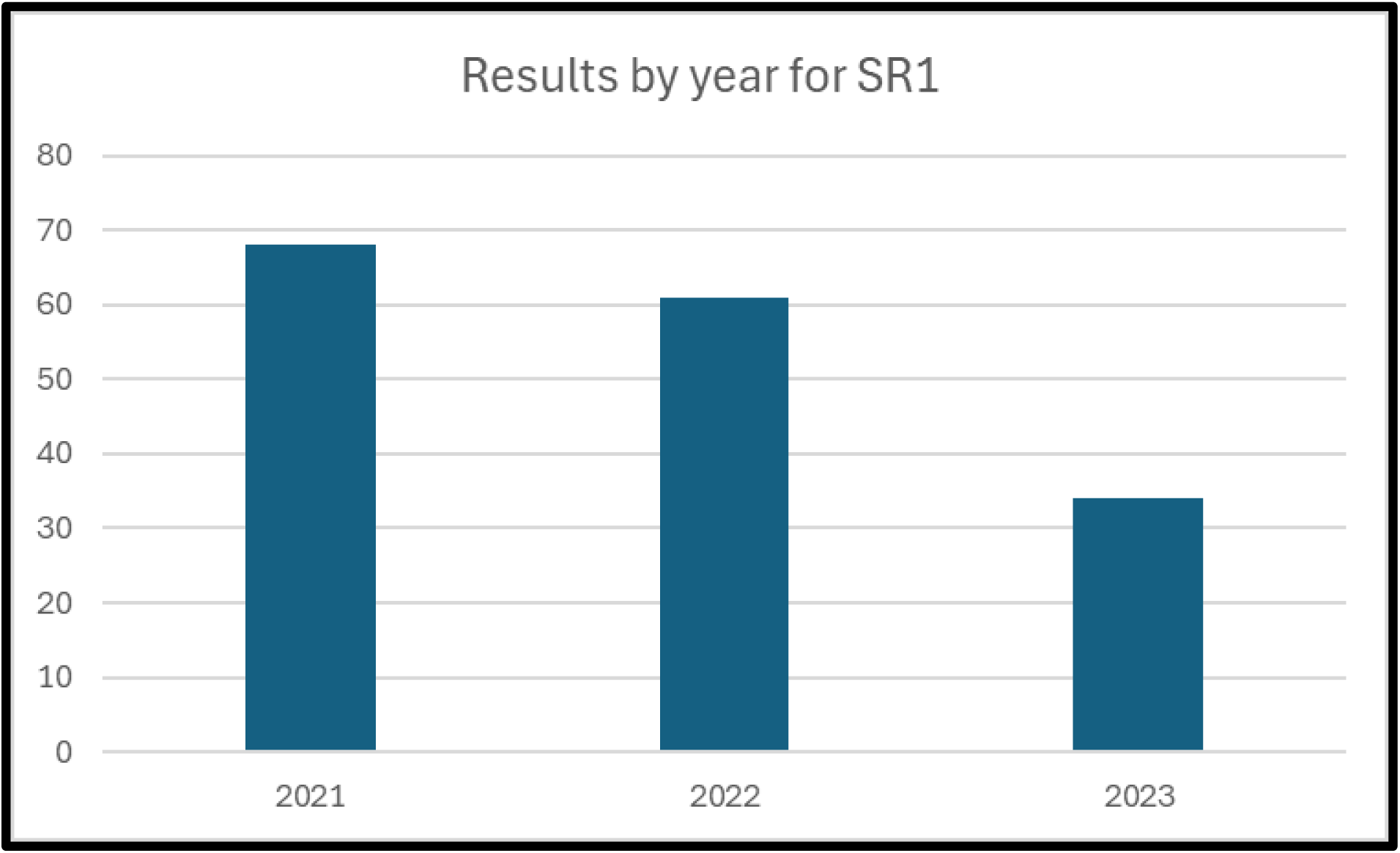
Number of articles returned by year for SR1. Please note that the literature considered for 2023 encompasses articles published until August 2023, and this may explain the quantitative discrepancy with respect to 2022 and 2021.

**Figure 4.**
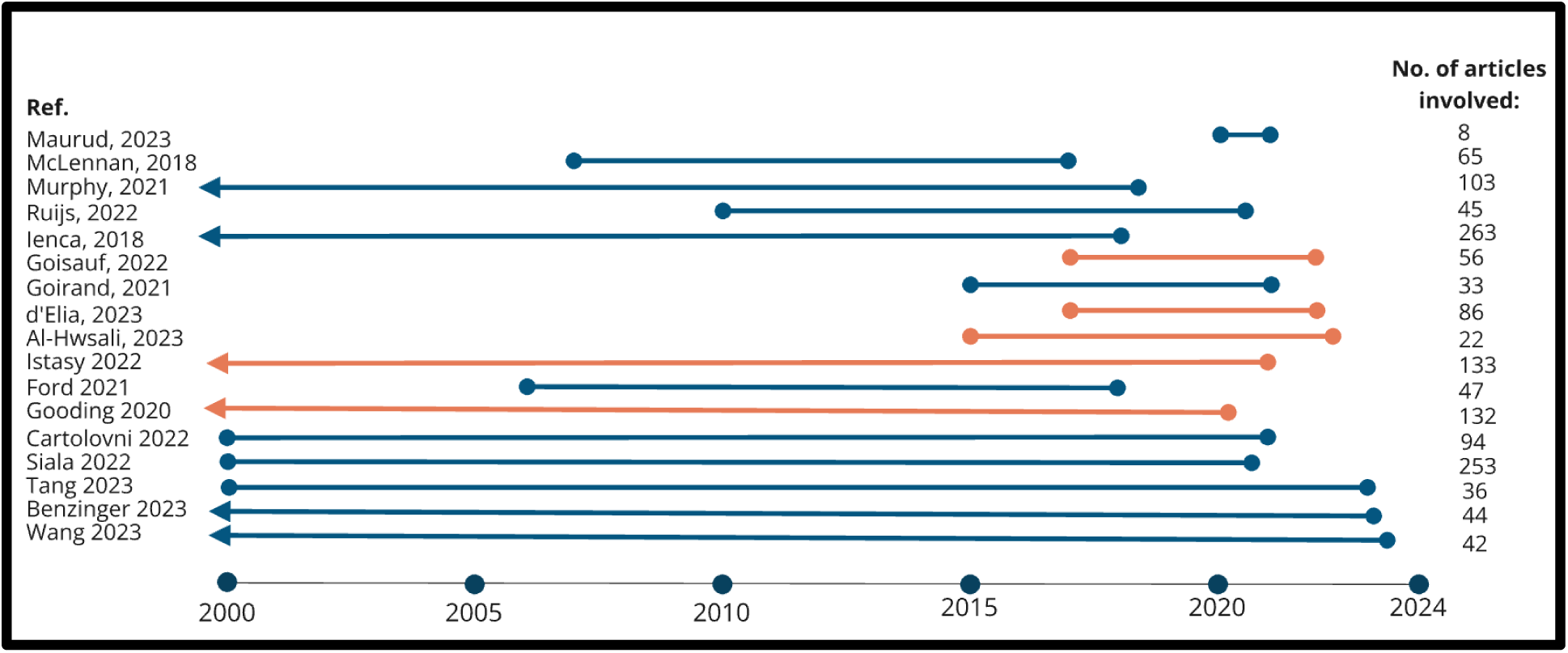
Included timeframe and number of articles identified in reviews found by SR2. Ref. – Reference of the review article, No. of articles included – Number of articles included on the reviews, Blue line – the review had a general scope in terms of field of medicine, Orange line – the review had a specific field of medicine as a scope, Arrow – the review had no minimum date set in their search sting

### 3.2 Methodologies of included papers

The majority of publications returned in SR1 were non-empirical studies (n = 142) while empirical studies were a minority (n = 23). This makes sense given that ethical consideration of novel, emerging technologies typically begins with philosophical analysis and only later progress to empirical studies as real world examples start to emerge which can then be studied. For SR2 all papers were necessarily systematic (scoping) reviews as this was a key inclusion criteria of our search. Of the empirical studies reported in SR1 a variety of common methodologies were employed, with the most frequently reported methodology being qualitative, semi-structured research interviews (see Figure 5).

**Figure 5.**
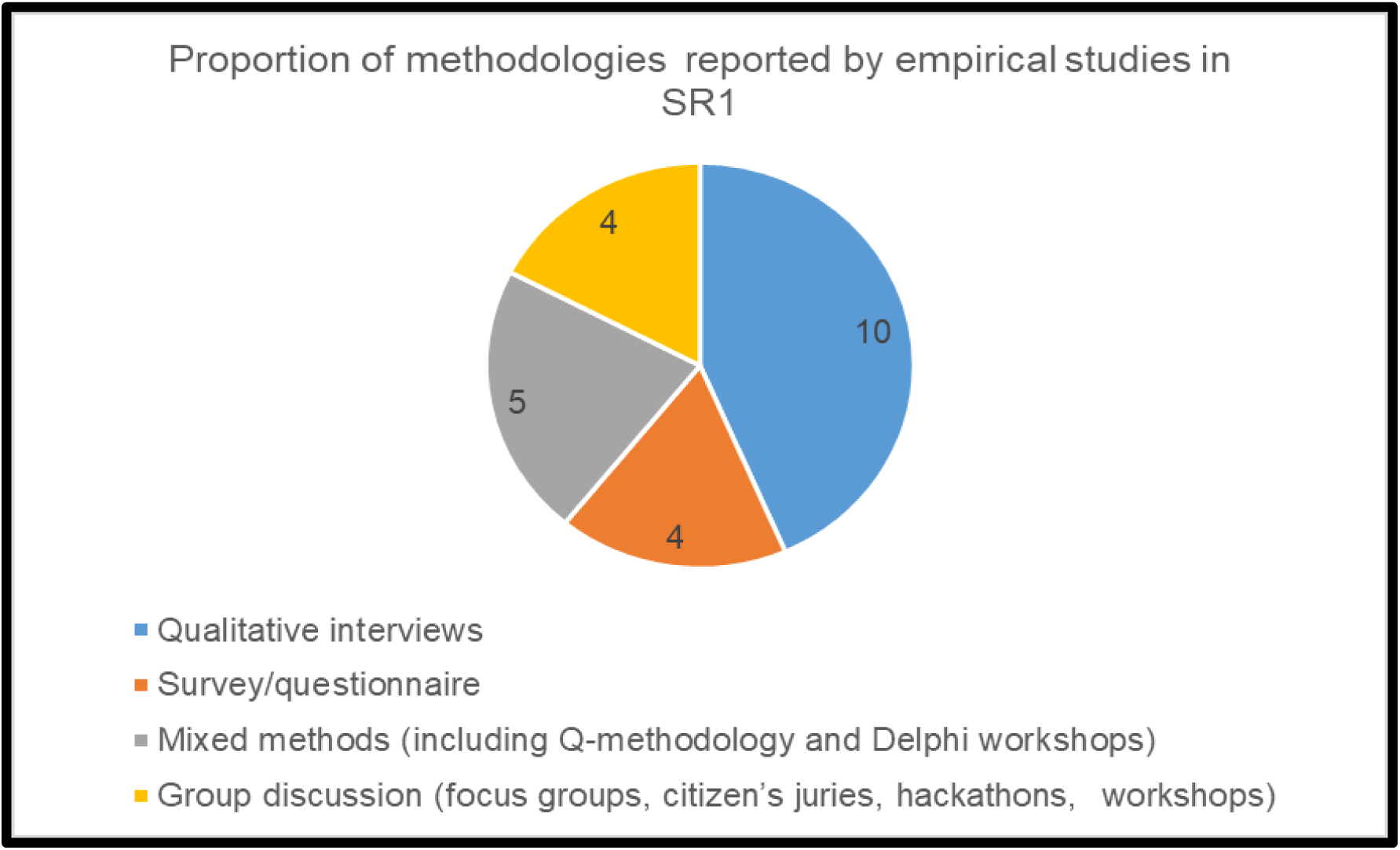
Proportion of methods reported by empirical studies in SR1

Of the empirical studies reported in SR1, the majority were conducted in a single country (n=15), some were conducted in multiple countries (n=6) and two reported no data on the location of the study (n=2). The most commonly reported geographic region for empirical studies was Europe only (n=9), then North America only (n=3), with additional papers reporting on studies from both European and North American countries in the form of a comparative analysis (n=3). Two studies were conducted in Asian and African countries only respectively (n=2), and a single study reported from Australia (n=1).(Figure 6).

**Figure 6.**
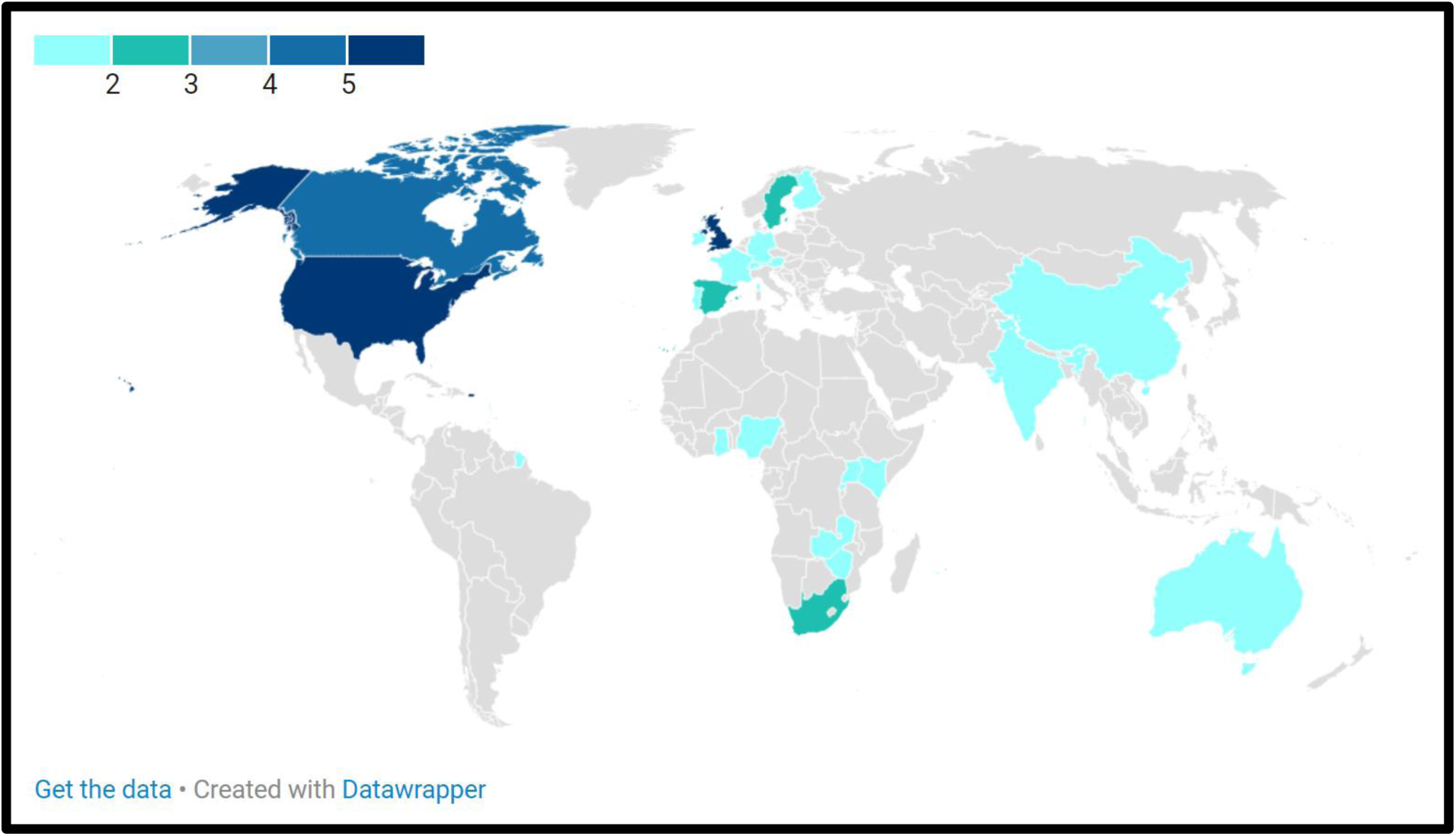
Map showing frequency of countries included in empirical studies

### 3.3 Audiences and subjects of study

The empirical studies in SR1 reported a variety of different subject groups enrolled in the study (see Figure 7). Mixed groups of different types of experts (and or lay representatives) were most commonly used, followed by studies with healthcare professionals (n=5) and patients (n=4). This makes sense given the interdisciplinary nature of AI and the healthcare setting.

**Figure 7.**
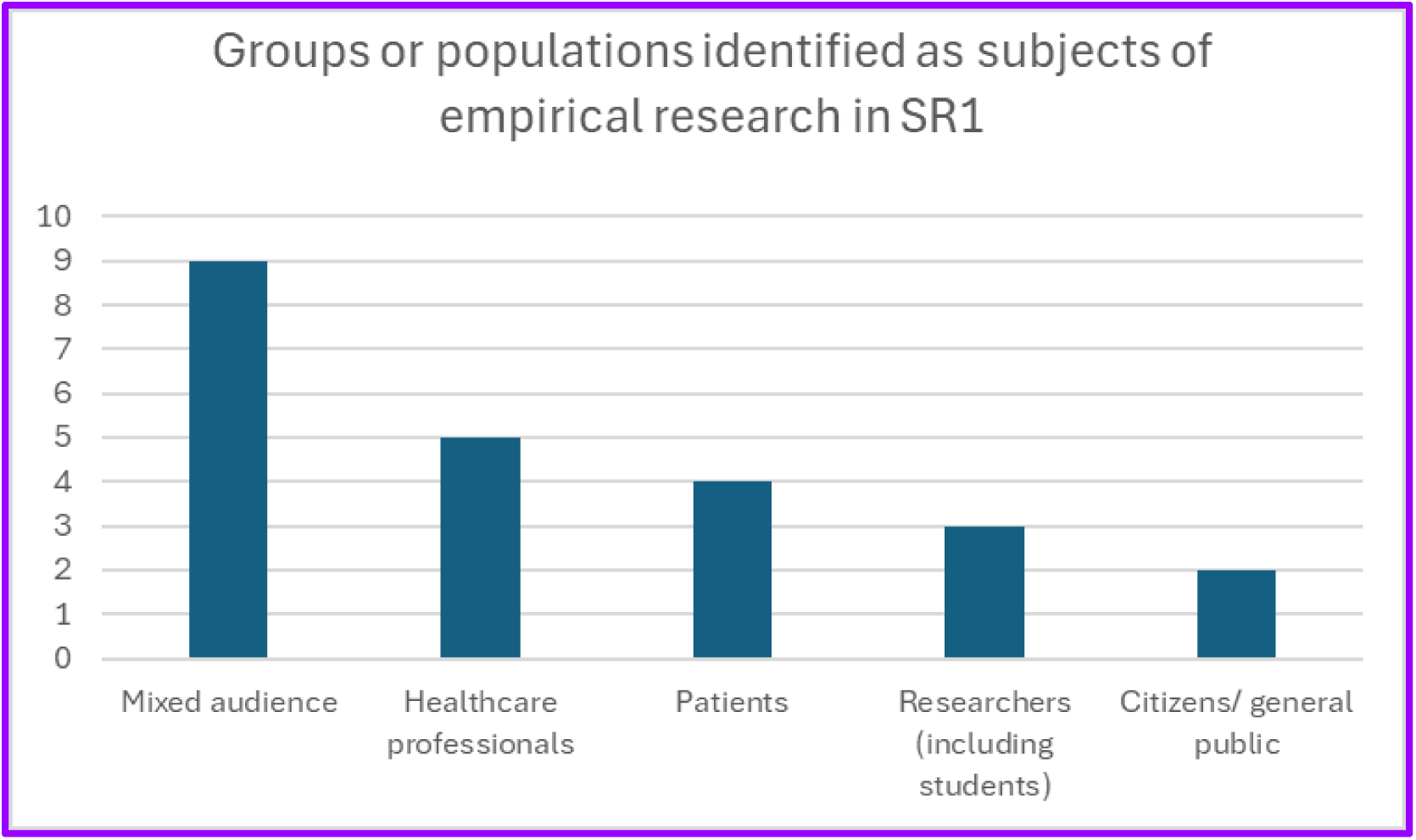
Groups or populations identified as subjects of empirical research in SR1

Where information was reported, a majority of empirical studies used hypothetical scenarios describing potential uses of AI in healthcare to elicit responses (n=5) rather than compiling experience based recommendations, suggesting that a good deal of AI ethics discussion remains anticipatory in nature. Nonetheless, overall most studies did not provide this information (n=12).

Audiences to which articles were directed also varied significantly. Articles were explicitly directed to patients in 23 cases; to healthcare professionals (including nurses and physicians) in 32 cases; to developers in 20 cases; to regulatory authorities or regulatory audiences in 14 cases; in 85 cases it was not specified. (see Figure 8).

**Figure 8.**
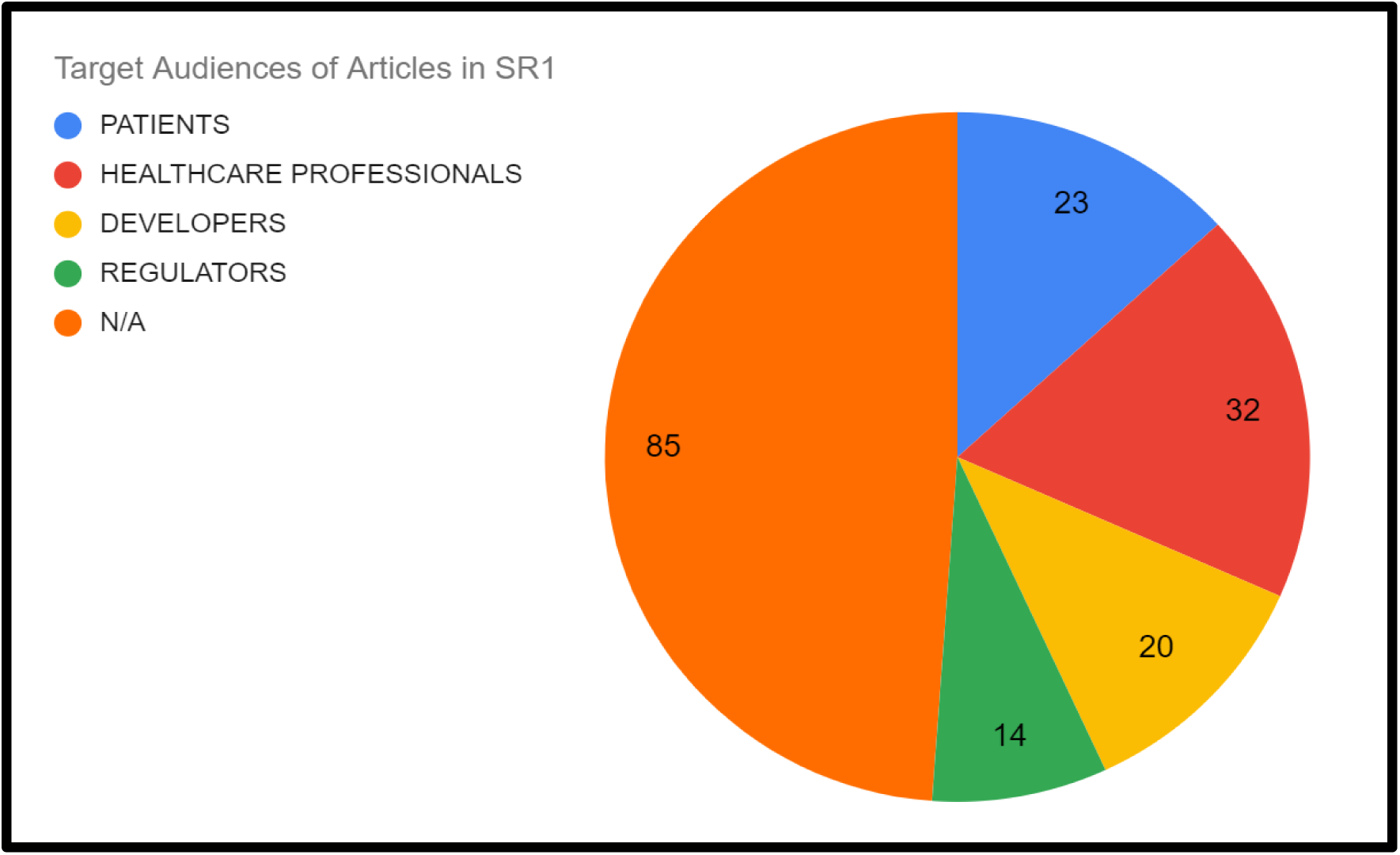
Proportion of articles directed to particular audiences in SR1

### 3.4 Medical domain and type of application of AI

Among the papers returned in SR1, the majority either did not specify a specific area of medicine or were considered to cover medicine in general (n = 78). Where a field or type of medicine was specified, the most commonly addressed area was mental health, broadly defined to include applications in psychology and psychiatry, dementia research et cetera (n=13), followed by a range of public health applications including covid-19 and HIV surveillance (n=8). There was a long ‘tail’ of medical domains that only received one or two papers, as the use of AI in niche or specialist areas of medicine typically attracted limited attention, with articles usually published in journals dedicated to those particular medical specialisms. The full range of medical domains reported, and their respective rates of occurrence in SR1, are displayed in Table 1.

**Table 1.** Range and frequency of medical domains in which ethical and social issue of AI were addressed in SR1.

| Medical field | Frequency |
| --- | --- |
| General/ not specified | 100 |
| Mental health and psychiatry | 11 |
| Public health | 9 |
| Radiology | 7 |
| Cardiology | 6 |
| Surgery | 4 |
| Neurology | 3 |
| Intensive care | 2 |
| Oncology | 2 |
| Pathology and laboratory medicine | 2 |
| Pediatrics | 2 |
| Personalised medicine | 2 |
| Randomized clinical trial | 2 |
| Other (one each: Dermatology, Disability, Emergency care, Gastroenterology, Gerontology, Gynecology, Incapacitated patients, Inpatient care, Nephrology, Orthopedics, Pulmonology, Ophthalmology) | 12 |
| Total | 164 |

Areas of medicine considered in the scoping review papers returned in SR2 were medicine in general (n = 12), public health (n = 1), dentistry (n = 1), primary care (n = 1), pharmacovigilance (n = 1), mental health (n = 2), radiology (n = 1), clinical research informatics (n = 1). In both SR1 and SR2, the majority of studies refer to AI broadly conceived (as to include most common types of algorithmic or Big Data technologies) or without specifying the type of AI (SR1 n= 70; SR2 n=18). In SR1, where an application type was specified, by far the most common type was diagnostic and treatment applications of AI (n = 60), with only very small numbers of studies falling into the categories of databases (n=3), conversational tools and chatbots (n=.3), patient engagement and adherence tools (n=2) and ‘other’ (n=10). The latter category included types of application such as LLMs, ‘digital twin’s et cetera In SR 2, two studies referred explicitly to Natural Language Programming as the type of AI addressed in the review while the others did not specify a type of application.

### 3.5 Ethical and societal themes addressed in SR1 and SR2

In both SR1 and SR2 a wide range of different ethical and societal issues were reported. SR1 utilised our *a priori* list of issues to classify the contents of each paper (see Appendix 3). Articles were coded to allow for multiple themes to be captured for each paper, reflecting the fact that some papers focused explicitly on one dimension such as ‘trust in AI systems’ while many others covered multiple issues pertaining to the ethical application of AI in healthcare. As a result, there are more codes reported for each arm of the scoping review than the total number of papers in either of SR1 and SR2. In SR1 the largest single category was the ‘other’ category for novel, unexpected or miscellaneous themes, where these are ‘novel’ or ‘unexpected’ with respect to widespread general AI ethics principles mentioned in the literature (Jobin et al 2019), as we show in the Discussion. The named issue categories that attracted the most frequent mention among papers in SR1 were ‘fairness/bias’, ‘privacy and data protection’ and ‘trust’, as evidenced by Figure 9.

**Figure 9.**
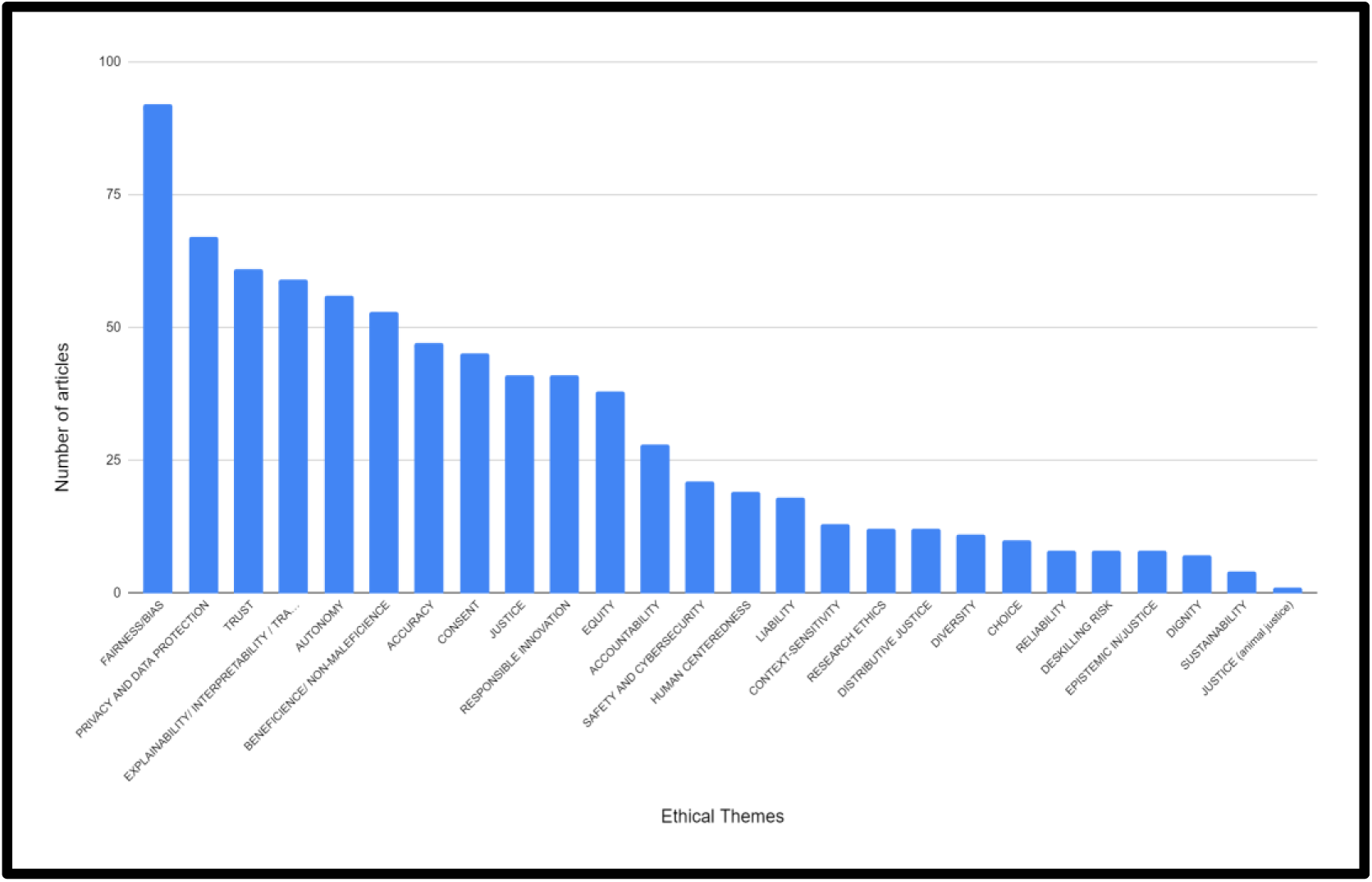
Ethical themes in SR1

A similar pattern was seen in SR2. As noted, SR2 did not use predefined categories, but reported the major ethical and societal issues reported by each scoping review using that article’s own terminology. However, since the use of terminology was not consistent across papers, we have aggregated similar terms such as ‘privacy’ and ‘confidentiality’ or ‘justice’, ‘social justice’ and ‘fairness’ into composite categories as displayed in Figure 9. The three most frequently reported categories in SR2 were ‘privacy and confidentiality’, ‘bias’ and ‘trust/trustworthiness’ as shown in Figure 10. In both SR1 and SR2 the next most frequently encountered category was ‘autonomy’, while ‘consent’ ‘justice’ and ‘transparency’ were also popular themes in both strands of the review. SR2 has a longer ‘tail’ of issues receiving only one or two mentions. Many of these issues would have been compiled into the ‘other’ category in SR1 which goes some way to explaining its high frequency. Notably, our *a priori* categories ‘research ethics’ ‘epistemic injustice’ and ‘responsible innovation’ did not occur in SR2, suggesting that these terms, and the concepts they represent, have not been used widely in the history of debates on the ethics of AI in healthcare.

**Figure 10.**
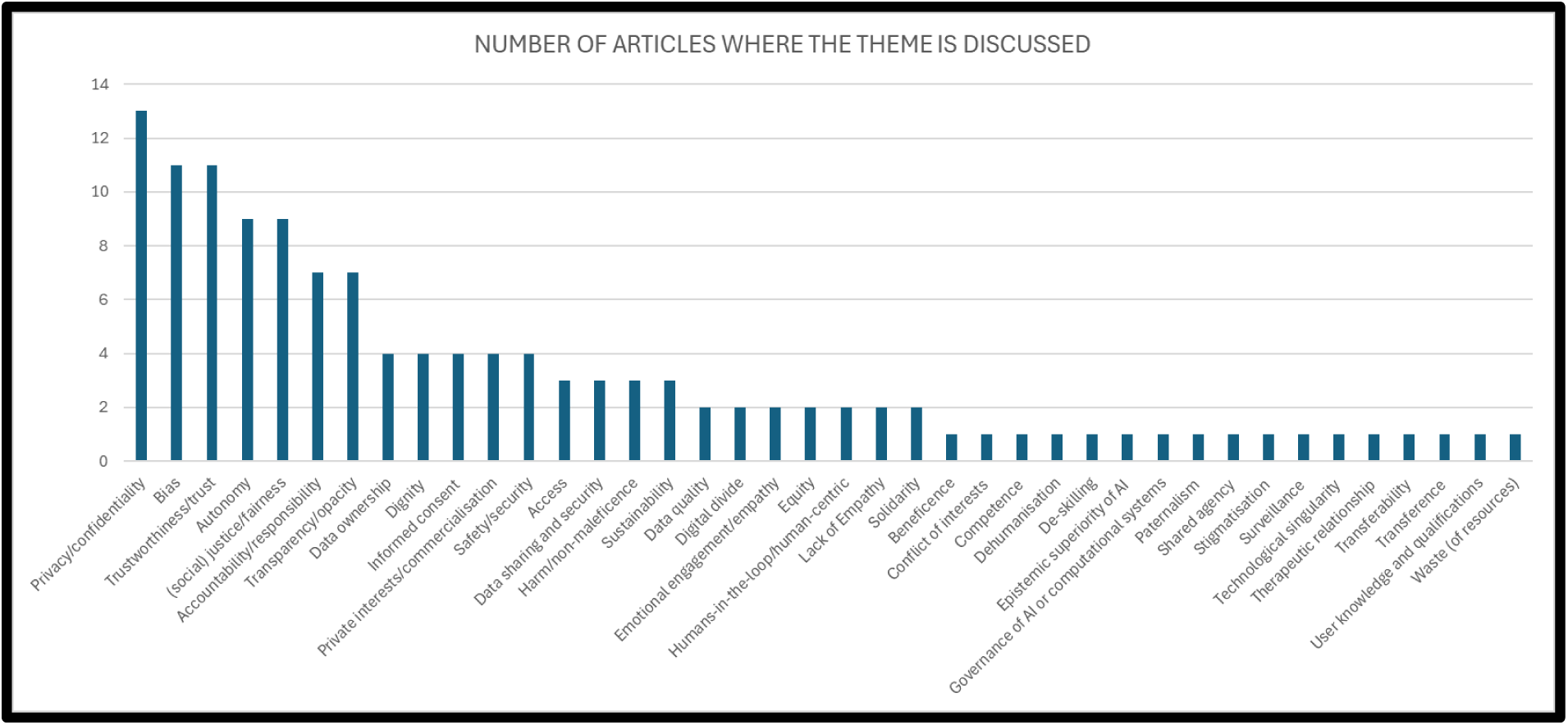
Ethical themes identified in SR2

A total of 114 entries were recorded in the ‘other’ category in the broad review (SR1). These covered a wide variety of terminology and phrasing and it was decided that this data needed further clarification to be of use. The full list of ‘other’ values reported in SR1 was reviewed by all three authors and each entry was either assigned as being a subclass of, or adding a particular nuance to, one of the existing categories of values reported in SR1 (as described in Section 2.4 above) or was recognised as belonging to *a novel category* not deployed in our list of a priori ethical and social issues used to chart data in SR1. The new categories derived from reanalysing the ‘other’ column were numerous. One example is the deskilling of medical professionals (Parviainen & Rantala 2022; Quinn et al 2021; Milton 2021), which can potentially harm patients in the long run, but it is not captured just by a vague concept like ‘maleficence’. Another example of a novel category is ‘human-centredness’, which captures the impact of medical AI tools on human relations (Milton 2021). The need for this new category has emerged in discussions on ‘care’, which plays a central role in a few articles (see, e.g., Quinn et al 2021), while as a notion it is typically neglected in the field of AI ethics. Ultimately ‘care’ was coded as a specific dimension of ‘human centredness’ in our analysis, because concerns over the quality of care are also related to how AI tools are going to influence care practices, where the ‘human’ dimension is central. A growing concern which is not covered in classic discussion of AI ethics is animal suffering - in one article (Popa et al 2021) AI is also seen as contributing to what we termed ‘animal justice’ due to its potential to reduce animal suffering by replacing experimental animals with digital models. Environmental and economic sustainability of AI is another important challenge raised by AI tools that has not traditionally fallen within the scope of ‘classical’ discussions of AI ethics, although it is now beginning to garner ethical attention (Samuel and Lucivero 2020; Lucivero 2024), and again warranted a *de novo* category as part of the coding of the ‘Other’ category of ethical and social issues. Another novel theme identified was a possible concern over the high cost of AI and how this could limit its use. This is an interesting issue, because typical discussions in AI ethics are about deliberately limiting the use of AI. This was also connected to issues about the digital divide which, quite strangely, are neglected in AI ethics (probably for the emphasis on limiting the use of AI, rather than promoting it). These elements were often captured in another *de novo* category Distributive Justice (see Table s8 in Appendix 6). Although properly recognized as a subset of justice concerns, we decided to create a specific separate code for Distributive Justice to capture a growing number of discussions about global access to AI and the global distribution of benefits and harms of AI, especially with respect to divides between the the global north and global south, as well as between wealth and poor populations within countries and regions (Spiegel et al 2021). The definitions of these new ethical and social themes are presented in Table s9 (Appendix 6). The meanings of these emergent categories and the nuances that the entries in the ‘other’ category added to discussions of more common principles of AI ethics such as privacy and bias are shown in Table s9, Appendix 6.

While we have not provided a similar analysis for SR2, there are significant hints that in existing scoping reviews, the typical way to understand AI ethics principles might have important limits, and that hyperspecialization (as we further discuss later) is a growing trend. For instance, D’Elia et al (2021), which has a short span (2017-2021), and it is on a narrow topic (i.e., primary care), address specific questions about the doctor-patient relationship. The nuances (e.g., participation, dehumanisation, agency of self-care) of this topic may not be easily covered by the general principles. Another aspect suggesting the increasing hyper-contextualisation of ethical and societal issues is the lack of homogeneity in reporting gaps. Depending on the context and research questions, different gaps were reported, where these gaps can hardly be reducible to a general and high-level discussion of AI ethics. For instance, in the context of dentistry (Favaretto et al 2020), there is a lack of discussion on issues raised by 3d imaging technologies, which are especially salient in dentistry, but they may not in other fields (e.g. mental health). Finally, where reviews analysed in SR2 reported trends that are in principle categorizable in terms of the general principles of AI ethics, they nonetheless add nuances that point to the limitations of using those principles for analysing ethical and societal issues in this variegated context. For instance, Li et al (2022) stress the importance of data privacy. However, privacy in that work is mostly seen from the angles of data ownership, stigmatisation, dignity, and well-being. These are rich concepts that are hardly reducible to blanket characterizations referring to, e.g., respect for autonomy or non-maleficence. This is to suggest that a discussion of data privacy that only mentions respect for autonomy or non-maleficence might miss important challenges and nuances that a discussion based on ownership, stigmatisation, dignity and well-being would not.

An important aspect of the literature of both SR1 and SR2 is the type of tradeoffs among societal and ethical values. By ‘tradeoff’ here we mean that articles have emphasised that the use of AI in medicine may promote certain values at the expense of others. A typical tradeoff often commented in the literature is that if one wants to have more accurate AI tools (where ‘accuracy’ is a value, in the sense of a desideratum of AI tools), then this often comes at the expense of interpretability/explainability. In other words, you cannot increase accuracy and interpretability/explainability at the same time. In SR1, accuracy vs explainability is widespread as noticed in (Liao 2023). Wehrens et al (2023) emphasise the importance of finding a balance between values that are perceived as typically in tradeoff, such as economic vs public values, data protection vs patient protection. Mascalzoni et al (2022) and Balthazar et al (2021) identify a tension between openness and privacy. Other examples of tradeoffs include fairness vs accuracy (Cohen et al 2022), regulation vs speed of innovation (Knoppers et al 2022). Another interesting tradeoff is between individual benefits vs group benefits. For instance, Stake et al (2022) notice that collecting sensitive data on a healthy child in an app might pose some privacy risks to the individual child, but at the same time the information may benefit certain groups. For instance, they mention that “children often continue to receive medications ‘off-label’ and the dosage is often based on the dosage for adults, as reliable data for children is lacking” (p 6). By collecting more data, these problems may be addressed more effectively for children collectively (i.e. as a group) even if this increases the identifiability and thereby reduces the privacy of individual paediatric patients. A number of scoping reviews analysed in SR2 also emphasised the existence of value tradeoffs. For instance, Goirand et al (2021) notice that beneficence can be compromised by fostering autonomy, and that in virtual bots for elderly care, trust may be compromised to ensure safety. They also point out that different dimensions of fairness are typically in contrast, as also widely known since the formulation of impossibility theorems for fair-ML (Kleinberg et al 2017). Another tradeoff mentioned here is the classic one of accuracy vs transparency (Goisauf et al 2022). Li et al (2022) comment on possible conflicts between different objectives of local healthcare facilities and the high-level policies dictated by existing hard and soft regulations, or between patients’ and medical professional and insurance companies. Reyes et al (2022) notice that lack of stakeholder engagements can lead to conflicts in the ethical and societal values to embed in AI tools.

### 3.6 Trends identified in SR2

A majority of papers in SR2 (n=12, 80%) reported some trend or pattern in the literature they reviewed. The remaining 40% of papers (n=8) did not present any identifiable discussion of a trend in findings. Of those papers where discussion of a trend was discernible, the most common type of trend was ‘issue based’. This means that what we consider a ‘trend’ in these cases was a reported prevalence of a particular ethical and/or societal issue in the literature covered by that specific scoping review. This reflects the fact that most, though not all, scoping reviews in SR2 sought to identify key ethical issues in particular areas, whether subdomains of medicine or associated with particular types of application. ‘Privacy’ was most often reported as the most frequently discussed issue or the issue which attracted the greatest amount of discussion, with ‘bias’ a close second. These trends are equivalents of the type of findings reported for this study in section 3.5. However they do not match exactly our ranking of the prevalence of issues found in either SR1 or SR2, partly because each scoping review asked a different research question and because, as reported in Table 4, there was significant variation in the time periods covered by each review and the volume of literature considered.

Other studies identified the issues the authors considered most important ( which was not always based on prevalence), for example Cartolovni et al (2022) state “the most critical social issue identified by our literature review is the impact of AI on the patient-physician relationship”. Some scoping reviews in SR2 also noted which issues were least discussed such as social justice (Al-Hwsali et al. 2023), explicability of AI outputs (Benzinger et al. 2023) and Ienca et al (2018) who observed that “issues of data ownership, group-level ethical harms, and the distinction between academic and commercial uses of big data, do not appear as ethical priorities”. Maurud, Henni and Moen (2023) examined the discourse on equity specifically, finding that the scope of discussion of health equity in clinical informatics had expanded from an early focus on race and ethnicity to encompass “diagnosis bias in rural populations, age, and gender or sex-specific bias”.

The remaining papers in SR2 in which any form of trend was discernible focused on ‘outcome’ based trends. These occur where the research question addressed by the scoping review set out to identify recommendations for best practice (e.g. for use of social media health data as investigated by Ford et al 2021) or most frequently discussed challenges and benefits to e.g. implementation research ethics review of AI in healthcare (Ienca et al 2018; Goirand et al. 2021). These trends pointed to findings such as the need for researchers to determine whether, and under which circumstances social media users should count as human subjects for research ethics oversight (Ford et al 2021), the risk of AI in primary care exacerbating health inequalities (d’Elia et al. 2022) or the major types of challenge facing ethical frameworks for AI (Goirand, et al. 2021).

### 3.7 Gaps identified in SR2

An important aspect of scoping reviews is their perception of things that need to be discussed, but that they could not find in the articles they analyse. Thirteen articles discussed gaps in the literature. The gaps identified are highly specific to the context of the different scoping reviews, and for this reason they are sometimes surprising. For instance, Al-Hwsali et al (2023) notice that in AI-based public health there is no accountability framework. Rather than a gap in the literature, they seem to refer to a lack of effort to create such a framework. This is surprising because there are plenty of discussions on accountability in AI, but apparently not in the specific context of public health at the time of that review. Another interesting result comes from Benzinger et al (2023). They point out that in AI-based clinical decision-making there are not many discussions on justice, and they think that this depends on the context dependence of justice, which makes it impossible to have an overarching concept of justice in place. In the context of radiology, Goisauf et al (2022) are concerned with the lack of attention on bias, and especially on gender bias. While issues of informed consent are typically at the forefront of discussions in biomedical ethics and AI ethics, Goording et al (2020) notice that when AI is used in the context of mental health, this is not as prominent as it should be, potentially due to concerns over mental health patients’ capacity to consent (see e.g. Skorburg, O’Doherty and Friesen 2024). In the context of dentistry, Favaretto et al (2020) notice a number of gaps, which are due to how recent ethical issues in AI dentistry are.

Other issues are less surprising, and seem to identify gaps in discussions on ethical and societal issues in medical AI more in general. For instance, Cartolovni et al (2022) are especially concerned with the lack of qualitative and quantitative studies of ethics of AI in the medical context. Ienca et al (2018) emphasise the importance of re-use of data, and data-subjects’ control of data. Istasy et al (2022) make a plea for more discussions on the importance of infrastructure and human resources in the analysis of ethical and societal issues raised by medical AI. Others notice that the lack of applicability of high-level principles to practices can impair the ethical dimension of AI tools, and so far this is not much discussed in the literature (Tang et al 2023).

## 4. DISCUSSION

Our data charting process and results suggest a number of interesting points worth discussing.

### 4.1 Ethical and Societal Issues and Their Connections with Epistemic Problems

One aspect that might not go unnoticed is that some themes that we have discussed are not strictly speaking limited to ‘ethical and societal issues’. For instance, accuracy and explainability are typically thought to be epistemic themes, where the former is about the performance metrics to evaluate algorithmic systems, while the second refers to a strategy one can use to inform discussions on the reliability of algorithmic systems. However, it has been also shown that such epistemic issues overlap significantly with ethical and societal issues. For instance, it is long known that choosing one performance metric rather than another is not just an epistemic issue: deciding whether to favor false positive at the expense or false negative (or vice versa) has important ethical ramifications because it can promote certain values rather than others (Biddle 2021), or in general the choice of the metrics itself can be motivated by appealing to certain values rather than others (Kraemer et al 2011). Explainability has a similar entanglement with ethical and societal issues. For instance, it has been recognized as a moral and epistemic principle at once (Floridi and Cowls 2019), as it is sometimes seen as providing legitimacy (or not) for automated decision making in highly-sensitive contexts. Even when it is not seen as providing legitimacy *per se* (Henin and Le Matyer 2022), it is difficult to deny that it can be seen as a necessary starting point for ethical notions such as contestability.

### 4.2 The Scope of Medical AI Ethics

A second important observation is that some ethical and societal themes go well beyond the scope of the ‘headlines’ of medical AI ethics. For instance, sustainability, environmental impact, and animal well-being are rarely mentioned in popular accounts of AI ‘disruptions’ in healthcare. Yet, our scoping reviews have identified interesting connections between these themes, and the use of AI in healthcare. This suggests a picture in which ethical and societal issues raised by AI cannot be neatly anticipated just by looking at the contexts, even though the way in which algorithmic systems interact with the specificities of contexts will result in unpredictable ethical and societal disruptions.

This second observation was especially remarkable in SR1. In this regard, SR1 is characterized by two aspects that are somewhat in tension. On the one hand, we started our analysis with a list of recurring *a priori* themes that we elaborated from our existing knowledge of ethical and societal issues in medical AI (see Appendix 3). These themes overlap significantly with lists of high-level ethical and societal principles that inform much of the debate on AI ethics in general (Jobin et al 2019), and hard regulation (Ratti and Graves 2025). In the AI ethics literature, it is typically thought that guidance on how to responsibly develop and deploy AI tools should be informed by five principles: beneficence, non-maleficence, justice, respect for autonomy, and explainability (Floridi and Cowl 2019). The articles in SR1 discuss at great length these themes we start with. This means that much of the literature on the ethical and societal issues raised by medical AI overlaps with classical discussions in AI ethics. This is reinforced by the finding that there is a strong overlap in the most frequently reported categories of ethical and societal issues across SR1 and SR2, such as privacy/confidentiality, bias, fairness, trust/trustworthiness, justice, consent and so on. This suggests these issues persist over time and constitute a defining ‘core’ of ethical and societal issues in healthcare. For instance, the most recurring themes identified in the articles fall under at least one of these principles, e.g., fairness/bias can be discussed as a justice issue, interpretability and transparency can be thought as issues pertaining explainability, privacy and data protection as pertaining to respect for autonomy, safety and cybersecurity as non-maleficence themes, and responsible innovation being entangled with beneficence.

On the other hand, in charting the data we needed to create an ‘Other’ column to capture those ethical and societal themes that did not obviously fall into any of the themes we start with (and hence into none of the five principles of AI ethics). Themes with such characteristics were so numerous that we have created another table to analyse them in more detail and establish whether they are indeed novel issues or just add nuances to known themes (see table s8, Appendix 6). In a significant number of cases, ethical and societal themes diverged significantly from known issues, and required the creation of new categories, such as deskilling, human-centredness, care, distributive justice, and others as we described in section 3.5. In other cases, unanticipated or hard to classify elements in the ‘Other’ ethical and social themes category were judged, during the recording of this data, to be interesting nuances or aspects of one of our a priori themes and added to the counts displayed in Figures 9 and 10. Again, the long ‘tail’ of categories reported in SR2 (see Figues 10) replicates and reinforces this pattern. Often these findings point to attempts to grapple with some of the practical challenges of developing, and using algorithms in healthcare. For example, several cases ultimately coded as issues of justice, related to the influence of the private sector in obtaining training data for AI tools and in shaping the design and future direction of AI research. But what both cases (de novo categories and nuances added to known categories) show, is that framing the discussions on traditional AI ethics principles is not always helpful, as the discussion is getting more and more fragmented due to the specificities of the way AI tools are designed and implemented in highly specific (and heterogeneous) contexts. SR1 shows this fragmentation in the many niche medical domains with only one or two papers dedicated to assessing the specific ethical and societal issues in that field or subfield, while SR2 revealed a number of narrow scoping reviews also focused on specific medical domains or use cases, such as public health, primary care, radiology, mental health, dentistry, indicating that some degree of fragmentation has been evident for some time. It is no surprise that high-level principles, in being general, become also uninformative to practitioners who are trying to navigate highly specific contexts, with highly specific epistemic and normative characteristics (Bezuidenhout and Ratti 2020).

This is evidence that a blanket application of the five principles in medicine in general does not do justice to the heterogeneity of ethical and societal issues raised by AI in specific contexts which typically entail multiple, overlapping ethical dimensions. This emerges also in reporting gaps. Depending on the context and research questions, different gaps were reported, where these gaps can hardly be reducible to a general and high-level discussion of AI ethics. But even when categorization happens through the lens of the typical principles of AI ethics, nuances are emphasized, and this suggests an inability of those general principles to make sense of the issues raised of AI systems, when they are implemented in specific contexts with their own normative constraints. This has a notable consequence. If we cannot anticipate ethical and societal issues by using the high-level principles, then what is needed going forward is more empirical ethics studies of real world applications of AI in healthcare (c.f Cartolovni et al 2022) and more reports of successful, partly successful and unsuccessful attempts to embed responsibility in medical AI at all stages from design to routine use and even disposal. This should lead to a richer discussion which, in turn, can be helpful to practitioners to design and implement medical AI systems that are indeed responsive to real issues and have a more concrete impact.

Overall these points of concordance between SR1 and SR2 suggest that the combination of broad and deep scoping reviews works to give a cumulative insight into both the sustained ‘core’ ethical and societal issues of AI in healthcare and its changing scope over time. As a final point of methodological reflection, it would have been interesting to run SR2 first and then derive the set of a priori categories of ethical issues from the findings of the deep review, and this is something we would consider for a future application of this method.

### 4.3 Tradeoffs in Values, Participatory Design, and Medical AI Ethicists

A third aspect that should be discussed is the importance of tradeoffs of ethical and societal values in designing and implementing AI systems in healthcare. As we have shown in 3.5, the inevitability of such tradeoffs is emphasized in the articles of both SR1 and SR2. The presence of tradeoffs points to the importance of including stakeholders in the design and implementation of AI systems to make sure that the values of those who are directly or indirectly impacted by AI systems are represented, or that the tradeoffs are discussed among the relevant parties. There have been generalist works in AI ethics capturing the importance of stakeholders participation (von Eschenbach 2021; Ratti and Graves 2025). In particular, there have been attempts to use value-sensitive design (Umbrello and van de Poel 2021) or socio-technical approaches (Fazelpour and De Arteaga 2022) to construct a method to increase participation of relevant stakeholders, so that values are traded-off as a result of a deliberation, rather than arbitrary and uninformed choices made by engineers. These works are ‘generalist’ in the sense that they do not account for the specificity of healthcare contexts, and hence more works to account for the diversity of healthcare contexts is required.

Related to this aspect, is the importance of redefining the role of medical and clinical ethicists in the context of medical AI. The surprising ethical and societal issues emerging from the interconnections between AI systems and highly-specific contexts point to the importance of carefully considering both these aspects. There have been criticisms of the excessive ‘black-boxing’ of technology inner functioning that takes place in the context of biomedical ethics (de Melo-Martin 2022). In the context of AI systems used in healthcare, a firm and solid grasp of the procedures used to build such systems, on how to evaluate their performance vis-a-vis clinical utility and validity, and an understanding of their limits and potentialities, should be a necessary requirement for clinical and medical ethicists. In other words, because of the priority of certain epistemic constraints over ethical conceptualizations (Grote 2024), then the training of ethicists should be changed accordingly.

### 4.4 Limitation of Our Analysis

The limitations of our research stem from the use of scoping review methodology rather than systematic literature review. While our approach aimed to identify the broadest databases spanning the fields of medicine, philosophy, and technology, a systematic review would typically aim to include all potentially relevant databases comprehensively. We deliberately designed our search terms to be broad; however, based on our findings, we recommend that future systematic reviews incorporate phrases related to principles (e.g., “responsibility,” “accuracy”) and terms indicating ethical themes (e.g., “opacity,” “explainable,” “black-box”) in their search strategies. For scoping reviews, we maintain that the terms used for our scoping reviews are adequate and align with its nature.

## 5 CONCLUSION

There has been an explosion of ethical and societal issues raised by AI in healthcare. This is caused by a ‘perfect storm’. Taken separately, AI and the medical context are known to raise challenging moral, social, and political issues. When AI and the medical context are put together, those issues are likely to be exacerbated. The volume of publications on this perfect storm is huge, and difficult to manage. For this reason, we think that a traditional scoping review is unlikely to provide a comprehensive picture of what is happening and of how these issues are conceptualised and addressed, and we propose a novel design to address this limitation. In this article, we have provided a two-pronged scoping review, which captures both very recent literature on ethical and societal issues raised by AI in healthcare (SR1), as well as past attempts to provide a comprehensive picture of those issues via scoping reviews (SR2). Overall the results of SR1 and SR2 can be shown to work in a complementary fashion to provide an account of the ‘core’ ethical and societal issues involves as well as indicating the move towards greater consideration of issues arising in particular niches, such as dentistry or public health, and to reflect in greater depth on the nuances and meaning of issues such as privacy in respect to AI in healthcare. Our results show that traditional methods in AI ethics based on high-level principles are unsuitable to analyze and integrate ethical and societal considerations in the design and implementation of AI tools in healthcare.

## 6. LIST OF ABBREVIATIONS

AI: Artificial Intelligence
ML: Machine Learning
SR1: Scoping Review 1
SR2: Scoping Review 2

## 7 DECLARATIONS

### 7.1 Acknowledgements and Funding

“This work was funded by the European Union under the Horizon Europe grant REALM 101095435. The views and opinions expressed are solely those of the author(s) and do not necessarily reflect those of the European Union. Neither the European Union nor the granting authority can be held responsible for them.”

### 7.2 Declaration of competing interests

IJ is a Board of Trustee at the European Patient Academy of Therapeutic Innovation (EUPATI) Foundation. Otherwise, all authors declare that they have no competing interests, whether financial, personal, or otherwise relevant to the content or choice of journal for this manuscript.

### 7.3 Data Availability statement

The authors confirm that the data supporting the findings of this study are available within the article and its supplementary materials. Raw data available from the corresponding author, [author initials], upon reasonable request.

### 7.4 Author’s contributions

ER has made substantial contributions to the conceptualization and design of the study; to the interpretation and analysis of data; and to the writing of the manuscript and appendixes.

MM contributed to the design of the study, interpretation, analysis and visualization of data, and to the writing and editing of the manuscript

IJ contributed to the design of the study, interpretation, analysis and visualization of data, and to the writing and editing of the manuscript

## Footnotes

2 We take a broad approach to defining ‘healthcare’, which we take to cover the application of knowledge and practices of biomedicine to human health in primary, secondary or tertiary care settings, and includes the fields of public health and mental health, but excludes complementary or alternative and non-Western medicine and broader domains such as wellness. For the purposes of this paper purposes of this paper, the terms ‘healthcare’, ‘medicine’ and ‘medical context’ are used interchangeably to refer to this domain.

3 There has of course been considerable interest in the ethics of AI in other areas of application as well, but this is beyond the scope of this paper.

4 A Guide to Knowledge Synthesis - CIHR (cihr-irsc.gc.ca), accessed 05/03/2024

5 This qualification was necessary to remove lower quality papers that define themselves as systematic or scoping reviews but which do not provide a clear account of their methodology and are more akin in practice to partial narrative or literature reviews.

6 For commentaries, abstracts were not available so decisions were made on title only.

7 Where this was reported the text was copied into Chart 2. In other cases it was necessary to infer the specific research question.

