## Appendix for "Ethical and Social Considerations of Applying Artificial Intelligence in Healthcare; a Two-Pronged Scoping Review"

**APPENDIXES**

### APPENDIX 1: SEARCH STRINGS FOR SR1 AND SR2

**Table s1: Search strings for broad review (SR1)**

| Database | Search string | Date run | No of results |
|---|---|---|---|
| Ovid Medline | 1 Artificial Intelligence/<br>2 artificial intelligence.tw.<br>3 computer vision.tw.<br>4 ethic*.tw.<br>5 exp Ethics/<br>6 2 and 4<br>7 1 or 2 or 3<br>8 4 or 5<br>9 7 and 8<br>10 limit 9 to yr="2021 -Current"<br>11 limit 10 to (yr="2021" and journal article) | 02/08/2023 | 411 |
| IEEE | ("All Metadata":"AI" OR "All Metadata":"artificial intelligence" OR "All Metadata":"big data" OR "All Metadata":"chatbot*" OR "All Metadata":"clinical decision* support system" OR "All Metadata":"conversational agent" OR "All Metadata":"deep learning" OR "All Metadata":"machine learning" OR "All Metadata":"software as a medical device" OR "All Metadata":"SaMD" OR "All Metadata":"digital assistant" OR "All Metadata":"virtual assistant" OR "All Metadata":"neural network") AND ("All Metadata":ethic* OR "All Metadata":moral* OR "All Metadata":responsible OR "All Metadata":"RRI") AND ("Document Title":"clinic*" OR "Document Title":health OR "Document Title":"healthcare" OR "Document Title":"digital health" OR "Document Title":*medic* OR "Document Title":"public health")<br>Filters Conferences; Journals;date range 2021-2023 | 01/08/2023 | 101 |
| Philpapers | ("Artificial Intelligence" OR "AI" OR "Big Data" OR "Chatbot" OR "large language model" OR "deep learning" OR "machine learning" OR "data science") AND ("clinic*" OR "health" OR "health care" OR "digital health" OR "medic*" OR "biomedic*" OR "public health") AND ("bias" OR "ethic*" OR "moral" OR "principle*")<br>Return works published between: 2021-2023 | 02/08/2023 | 33 |
| WoS | Topic = "AI" OR "artificial intelligence" OR "big data" OR "chatbot*" OR "clinical decision* support system" OR "conversational agent" OR "deep learning" OR "machine learning" OR "software as a medical device" OR "SaMD" OR "digital assistant" OR "virtual assistant" OR "neural network" | 01/08/2023 | 364 |

| | | | |
|---|---|---|---|
| | AND Topic = "clinic*" OR health OR "healthcare" OR "digital health" OR *medic* OR "public health"<br>AND Title = ethic* OR moral* OR responsible OR "RRI"<br>AND Index date = 2021-01-01 to 2023-07-13<br>Refined by<br>ISI core collection only<br>Languages: English<br>Document type: Article | | |
| | | **TOTAL** | **909** |

**Table s2: Search strings for deep review (SR2)**

| Database | Search string | Date run | No of results |
|---|---|---|---|
| Ovid Medline | 1. Artificial Intelligence/<br>2. artificial intelligence.tw.<br>3. computer vision.tw.<br>4. ethic*.tw.<br>5. exp Ethics/<br>6. scoping review.mp. [mp=title, abstract]<br>7. 1 or 2 or 3<br>8. 4 or 5<br>9. 6 and 7 and 8<br>10. limit 9 to yr="2014 -Current" | 20/03/2024 | 133 |
| IEEE | ("All Metadata":"AI" OR "All Metadata":"artificial intelligence" OR "All Metadata":"big data" OR "All Metadata":"chatbot*" OR "All Metadata":"clinical decision* support system" OR "All Metadata":"conversational agent" OR "All Metadata":"deep learning" OR "All Metadata":"machine learning" OR "All Metadata":"software as a medical device" OR "All Metadata":"SaMD" OR "All Metadata":"digital assistant" OR "All Metadata":"virtual assistant" OR "All Metadata":"neural network") AND ("All Metadata":ethic* OR "All Metadata": moral* OR "All Metadata":responsible OR "All Metadata":"RRI") AND ("All Metadata":"clinic*" OR "All Metadata":health OR "All Metadata":"healthcare" OR "All Metadata":"digital health" OR "All Metadata":*medic* OR "All Metadata":"public health") AND ("Document Title":scoping OR "Document Title":systematic AND "Document Title":review)<br>Filters: date reggae 2014-2024 | 18/03/2024 | 12 |

| | | | |
|---|---|---|---|
| WoS | Topic = "AI" OR "artificial intelligence" OR "big data" OR "chatbot*" OR "clinical decision* support system" OR "conversational agent" OR "deep learning" OR "machine learning" OR "software as a medical device" OR "SaMD" OR "digital assistant" OR "virtual assistant" OR "neural network"<br>AND Topic = "clinic*" OR health OR "healthcare" OR "digital health" OR *medic* OR "public health"<br>AND Topic = ethic* OR moral* OR responsible OR "RRI"<br>AND Title = scoping OR systematic AND review<br>AND Index date = 2024-02-29 to 2014-01-01<br>Refined by<br>ISI core collection only<br>Languages: English<br>Document type: Review | 18/03/2024 | 202 |
| | | **TOTAL** | **347** |

**APPENDIX 2: DATA CHART FOR SR1**

(available as a separate excel file)

### APPENDIX 3: DEFINITIONS OF ETHICAL AND SOCIETAL THEMES EMPLOYED IN CHARTING DATA FOR SR1

**Table s3: Ethical and societal Themes and their definitions employed in charting data for SR1**

| Ethical or societal theme | Consensus definition employed in charting SR1 | Notes |
|---|---|---|
| **Accountability** | If the article discusses the broad problem of who is accountable for the decisions made in a context where AI tools are used | |
| **Accuracy** | If the articles makes the connection between ethical and societal issues, and the AI goal of making predictions that are as accurate as possible (in contrast to, for instance, interpretability) | |
| **Autonomy** | If the article discusses the extent to which AI tools in medicine impinges on the autonomy of patients or of medical professionals. | Autonomy is broader than, and distinct from 'Choice' in that autonomy relates to peoples' capacities to make decisions and to choose courses of actions, which is broader than the choice of whether or not to be algorithmically processed or the right to contest the outcome of an AI decision |
| **Beneficence/ Non-maleficence** | If the article discusses the extent to which AI tools in medicine can either do harm or be beneficial | |

| | | |
|---|---|---|
| **Choice** | If paper discusses whether users (patients/healthcare professionals/other) should have, or are entitled to be offered, a choice about i) whether or not an AI is used to process their data or ii) whether or not to accept the recommendation of an AI regarding diagnosis/ prognosis/ intervention etc. | |
| **Consent** | If the paper discusses informed consent to algorithmic processing of data for any purpose | |
| **Context Sensitivity** | If the paper discusses the ethical importance of recognising some or all of the specificities of place, cultural values, practices, knowledges, beliefs, politics, social organisation, meanings, historical situatedness, individual capabilities etc. when considering the design, implementation and/or evaluation of the functioning of AI | |
| **Diversity** | If the paper discusses the need for AI to be trained on a representative/inclusive range of data covering e.g. ethnicity/ gender/ age etc | |
| **Epistemic Injustice** | If the article discusses to what extent the use of AI tools in medicine can be an obstacle to activities that are distinctively epistemic – when someone is wronged in her capacity as a knower | Not discussed much in the literature but was included as it is of interest to one or more authors. |
| **Equity** | If the paper discusses issues of inequality produced or exacerbated by AI tools or applications of AI to redress disparities, on the principle of equity (i.e. allocating greatest resource to those with greatest need). | |

| | | |
|---|---|---|
| **Explainability/ Transparency/ Interpretability** | If the paper discusses whether or not a human-understandable justification or rationale needs to be provided for decisions or recommendations made by an AI or if the paper discusses the value of making sure everyone involved knows i) when an AI tool is being used and ii) having access to an explanation of how or why that AI makes a particular decision | |
| **Fairness/ Bias** | If the article discusses issues of algorithmic biases in the design or implementation of AI tools in medicine or issues of fairness (e.g. in treatment outcomes) not directly flagged by the authors as Justice or Equity | |
| **Justice** | Related to 'Fairness' but also includes issues of social justice i.e. inclusion of/ outcomes for marginalised groups. | Distinction between 'Fairness' and 'Justice' is in large part made on the basis of whichever terms the authors of the paper being charted employ to frame their arguments. |
| **Liability** | If the paper discusses who is or should be held legally liable if something goes wrong in cases where an AI has input to, or makes recommendations in aid of clinical decision making, or how to assess said liability, and/or the impact uncertainty about liability might have on medical practice/ patient outcomes | Liability code refers primarily to the context of legal liability/ medical negligence suits, whereas the Accountability theme is broader and not necessarily legalistic and includes personal, professional and institutional responsibility. |

| | | |
|---|---|---|
| **Privacy and Data Protection** | If the article discusses issues of privacy or data protection explicitly | Code is for normative discussions of privacy and/or data protection. Purely technical discussions were excluded. |
| **Responsible Innovation/ RRI** | If the paper discusses i) the needs for developers or hospitals to consider and/or address the wider societal consequences of using that particular AI tool or ii) the ethical desirability of consulting various different stakeholders and integrating their concerns/ values/ needs into the development and/or implementation of the AI tool | |
| **Research Ethics** | If the article discusses ethical issues explicitly connected to the use of AI tools in pharmaceutical or clinical research including discussions of how to conduct IRB/REC oversight of AI research in medicine/healthcare | |
| **Safety and Cybersecurity** | Covers all normative discussions of patient safety, safety of subjects of algorithmic processing, and cybersecurity of AI infrastructure | Cybersecurity was a minor aspect of this category in practice. |
| **Trust** | If the paper discusses the importance of trust or trustworthiness in relation to AI tools/AI developers. I ticked this category regardless of whether it was the trust of patients, the public, physicians, regulators or any other party that was being discussed | |

| | | |
|---|---|---|
| **Other** | Anything that did not clearly belong in any of the above categories. | Entries in the 'Other' category were subsequently reviewed and either classified as a nuance to or special case of one of the existing themes in this table or recognised as a new category and detailed in Appendix 6 |

**APPENDIX 4: DATA CHART FOR SR2**

(available as a separate excel file)

**APPENDIX 5: DATE RANGE, RESEARCH QUESTION AND VOLUME OF RESULTS RETURNED FOR SCOPING REVIEWS IN SR2**

**Table s4: Date range, research question, and volume of results for scoping reviews returned in SR2.**

| Scoping review | Research question | Start date | End date | No of results |
|---|---|---|---|---|
| Al-Hwsali et al. 2023 | Which are the ethical and legal concepts related to AI in the public health domain? | 2015 | 2022 | 22 |
| Benzinger, et al. (2023) | What are the implications of supporting ethical decision-support with AI? | Not given but in date of earliest paper in review is 2007 | Not given but date of latest paper is 2022 | 44 |
| Cartolovni, et al. (2022)* | Which ELSI are most reported for AI-based medical decision support tools? | N/a | 2020 | 94 |
| d'Elia, A., et al. (2022) | What research currently exists on the effect of AI on primary care equity? | 2017 | 2021 | 86 |
| Ford, et al. (2021). | What are best ethical practices for NLP researchers using social media and patient forum data for health research? | 2006 | 2018 | 47 |
| Goirand, et al. (2021) | What ethics frameworks have been implemented in AI applications in healthcare? | 2015 | 2020 | 33 |
| Goisauf, and Abadia 2022 | What types of ethical issues are raised by the use of AI in medicine, and how are these issues tackled especially in radiology? | 2017 | 2021 | 56 |
| Gooding and Kariotis (2020)* | In what ways are algorithmic and data-driven technologies being used in the mental health context?<br>How and to what extent are issues of law and ethics being addressed in these studies? | N/a | N/a | 132 |
| Ienca et al (2018)* | To identify the promises and challenges of health-related big data research that have relevance for Ethics Review Committees | n/a | 2017 | 263 |

| | | | | |
|---|---|---|---|---|
| Istasy et al (2022) | What are the current and potential impacts of AI applications on health equity in oncology? | 1946 | 2020 | 133 |
| Li, Ruijs, and Lu (2022). | (1) what are the ethical issues related to AI in healthcare, and (2) what are the ethical strategies related to AI in healthcare? | 2010 | 2020 | 45 |
| Maurud, Henni, and Moen (2023) | What ways health equity has been promoted in clinical research informatics with patient implications | 2021 | 2021 | 8 |
| McLennan et al. (2018). | To determine systematically the full spectrum of ethical issues that are raised for stakeholders in a Learning Healthcare System | 2007 | 2015 | 65 |
| Murphy et al (2021)* | What ethical issues have been identified in relation to AI in the field of health, including from a global health perspective? | N/a | 2018 | 103 |
| Siala, and Wang. (2022) | To assess and bridge the gap between the theoretical frameworks of AI ethics and the actual stated worries and concerns of present and future stakeholders of medical AI. | 2000 | 2020 | 253 |
| Tang, Li and Fantus. (2023). | To examine the similarities and differences in the understanding of fairness, explore influencing factors, and investigate potential measures to implement fairness in medical AI across English and Chinese literature | 2000 | 2022 | 36 |
| Wang, et al (2023)* | To garner more profound understanding about responsible AI applications in healthcare | N/a | 2023 | 42 |

The timeframes also varied widely, from a single year to 74 years or more. Searches, marked with a '*' in the table, did not select a start date but opted to commence from the earliest available data held by each database. These searches combined findings from several databases each with different, and not always specified, dates when records commenced, which is why no date range is given for these searches.

**APPENDIX 6 ANALYSIS OF MATERIAL CODED AS 'OTHER' ETHICAL AND SOCIAL ISSUES IN SR1**

**Table s5 Assessment and classification of all entries in the 'Other' ethical and social issues in SR1**

| Ethical or societal issues classified as 'Other' in SR1 | Final categorisation and charting | Rationale |
|---|---|---|
| **Accessibility** | Fairness/Justice | Context was economic availability of AI tools in healthcare, so this was considered a dimension of the existing categories of Fairness and Justice |
| **Accessibility to global populations/ unequal access between global north and south** | Distributive Justice | New category: see table 9 below: see table 9 below: see table 9 below |
| **Accountability of designers and developers** | Accountability | A dimension of the existing theme |
| **Adequate reporting** | Transparency | A dimension of the existing theme |
| **AI may become standard of care and reduce physician judgement** | Deskilling Risk | New category: see table 9 below |
| **AI replacing medical doctors might increase the social withdrawal of psychiatric patients** | Human Centredness | New category: see table 9 below |
| **Algorithms for aesthetic surgery could impose western norms of beauty on non-western cosmetic surgery patients** | Fairness/Bias | A dimension of or nuance to the existing theme |
| **Benefit to science** | Beneficence | A dimension of or nuance to the existing theme |
| **Benefit to society** | Beneficence | A dimension of or nuance to the existing theme |

| | | |
|---|---|---|
| **Bodily integrity** | Non-maleficence | Avoiding harm to bodily integrity falls within the remit of the existing theme |
| **Bycatching** | Privacy and Data Protection | Terms used to describe instances when AI monitoring for example behaviour or speech collects data beyond what is needed for diagnosis/ detection and that may have unintended consequences |
| **Care (as a humanistic value)** | Human Centredness | |
| **Challenges to personal identity** | Human Centredness | |
| **Complexity** | Explainability | Complexity as a challenge to explainability/ transparency |
| **Concern that high cost of AI could limit its use** | Distributive Justice | |
| **Confidentiality** | Privacy | A dimension of the existing theme |
| **Conflict of interest** | Transparency | A dimension of the existing theme |
| **Consent** | Consent | Special case discussing the inadequacy of individualised Western model of consent for countries where community consent is more culturally relevant |
| **Contestability** | Transparency/Explainability | Contestability is about transparency/ interpretability/ explainability because it is only possible to contest a decision that can be understood or explained |

| | | |
|---|---|---|
| **Contextual integrity** | Context Sensitivity | Relates to issue of context of data use being different from context of data collection which can be disconcerting or distressing for data subjects. Also relates to privacy but is an explanation of how privacy relates to context. |
| **Data quality** | Accuracy | A dimension of the existing theme |
| **Data representative to target populations** | Fairness/Bias | Unrepresentative data can lead to biased outcomes. |
| **Datafication ("Datafication might force mental health professionals to translate patient information patient into a pre-configurated scheme, thus reducing individual characteristics to standardized categories")** | Human Centredness | |
| **Decolonialisation (including data).** | Distributive Justice | |
| **Deskilling risk** | Deskilling Risk | |
| **Diagnosis as interpretive, distributed exercise of expertise where AI can be a partner but should not determine outcome** | Autonomy | A dimension of or nuance to the existing theme |
| **Diagnostic AI may erode physician's practical wisdom in diagnosing** | Deskilling Risk | |

| | | |
|---|---|---|
| **Digital divide** | Equity | Relates to the risk of AI exacerbating existing inequalities due to uneven access to digital systems |
| **Dignity (including moral dignity)** | New category: see table 9 below | |
| **Diminished empathy through lack of contact with patients** | Deskilling Risk | |
| **Doing the right thing' by default.** | Beneficence | A dimension of or nuance to the existing theme |
| **Effect of using AI on the therapeutic relationship** | Human Centredness | |
| **Effect on health-care employer–employee relationships** | Human Centredness | |
| **Effect on patient-clinician relationship** | Human Centredness | |
| **Effectiveness.** | Accuracy | Effectiveness of AI depends on accuracy which is broader than representativeness of data |
| **Environmental impact.** | Sustainability | New category: see table 9 below |
| **Epistemic authority (when the reports of the individual are in conflict with the sensory data)** | Epistemic Injustice | Relates to the issue of whose claims should be heard |
| **Equal and fair access for citizens to the debate in the public sphere, not monopolised by academics, companies, regulators etc.** | Responsible Innovation | This is about stakeholder participation which is a key aspect of RI |

| | | |
|---|---|---|
| **Ethics of quantification- focus on measuring individual behaviours occludes social, environmental & other determinants of health** | Research Ethics | This dimension relates to ethics in design of algorithms in ways that implement a narrow definition of care |
| **Excessive reliance on mortality predictions may inhibit use of experimental therapies and learning by doing.** | Autonomy | A nuance to general discussions of autonomy focusing on how AI may inhibit physicians' capacity to innovate |
| **Freedom.** | Autonomy | A dimension of or nuance to the existing theme |
| **Generalisability.** | Bias | Relates to representativeness or otherwise of data and risk of bias |
| **Hidden labour of wearable sensors for diagnosis.** | Choice | Any requirement to wear sensors to generate data as a requirement to access care would be imposition on freedom of choice. |
| **High costs/ difficulty of integration** | Distributive Justice | |
| **Human in the loop.** | Autonomy | This frequently used phrase relates ultimately to opportunities for human decision making in Algorithmic contexts |
| **Human-centredness** | Human Centredness | |
| **Impact on medical jobs and morale.** | Human Centredness | |
| **Impact on vulnerable groups** | Equity/Justice | A dimension of the existing theme |

| | | |
|---|---|---|
| **Importance of local stakeholders** | Responsible Innovation | This is about stakeholder participation which is a key aspect of RI |
| **Inability of data based digital twins to think, feel or make choices** | Human Centredness | |
| **Inadequate regulation, expertise and infrastructure in Low and Middle Income Countries** | Distributive Justice | |
| **Inclusion** | Diversity | A dimension of the existing theme |
| **Inclusivity** | Fairness/Bias | Inclusivity in context of having representative data sets |
| **Individual informed consent cannot resolve community-level risk;** | Non-maleficence | A nuance to non-maleficence linking discussions of individual and group or community level harms |
| **Inequality** | Fairness/Bias | AQ brioad term that also relates to equity and/or justice. |
| **Infantilisation of users** | Autonomy | Context is AI might reduce human agency due to dependence on algorithms |
| **Integrity understood as 'the elaboration of respectful procedures for settling on social action despite the stand-off'** | Responsible Innovation | A dimension of RI related to stakeholder/ community engagement which is likely to produce disagreements |
| **Justifiability** | Transparency/ Explainability | Decision cannot be justified if it cannot be interpreted or explained |
| **Keeping promises** | Accountability | Of both AI and AI developers |

| | | |
|---|---|---|
| **Loss of critical practical nursing skills** | Deskilling Risk | |
| **Loss of human interaction** | Human Centredness | |
| **Loyalty** | Accuracy | In context 'loyalty' referred to reliability of algorithms in context of design parameters- do they do what they are expected and intended to do. |
| **Neutrality** | Fairness/Bias | Nuance to existing theme. Neutrality implies non-discrimination |
| **Opacity** | Explainability/ Transparency | Synonym for/ dimension of existing theme |
| **Open science/sharing data** | Transparency | A dimension of the existing theme |
| **Openness** | Transparency | A dimension of the existing theme |
| **Over-reliance dependence on AI** | Deskilling Risk | |
| **Ownership** | Transparency | Not data transparency but transparency in the context of conflict of interest arising from commercial ownership of AI tools |
| **Participation** | Consent | Context how medical professionals should explain the processes of diagnosis and treatment involving AI and gain adequate informed consent before enrolling patients in research or care |

| | | |
|---|---|---|
| **Paternalism** | Autonomy | Algorithmic paternalism could inhibit capacity of both doctors and patients to act |
| **Politics of innovation and governance.** | Responsible Innovation | Dimension of existing theme |
| **Possibility for citizens to contribute by providing/ selling data.** | Responsible Innovation / Distributive Justice | Relates to need for adequate social oversight and governance of such contributions and need to maintain access (e.g. to those who cannot contribute) |
| **Possibility for less invasive consultations.** | Beneficence | Nuance to existing theme in the form of a specific benefit or form of harm reduction |
| **Potential for increased friction between human doctor and patient if AI is wrong** | Non-maleficence | Increased 'friction' is harm to the physician-patient relationship |
| **potential for negative impact on medical staff and working environment,** | Human Centeredness | |
| **Potential to blur boundary of end of life** | Human Centeredness | |
| **Potential to reduce animal suffering** | Animal Justice | New category: see table 9 below |
| **Precedence of human values in decision making** | Human Centeredness | |
| **Private sector AI may favour profit maximisation over public good** | Justice | A nuance to the existing theme |
| **Private sector control of research may shape research direction** | Justice | Relates to the above, as private interests may not align with public good |

| | | |
|---|---|---|
| **Private sector led screening could foster overdiagnosis** | Non-maleficence | Over diagnosis, by definition is a form of harm |
| **Regulation/Governance** | Responsible Innovation | Appropriate and adequate governance is an aspect of socially responsible innovation |
| **Reliability** | Accuracy | Context of claims that AI functioning properly is more important than trust in a medical context |
| **Reproducibility** | Accuracy | A nuance to or dimension of the existing theme. |
| **Respect** | Autonomy | In the sense of respect for persons and personal autonomy. |
| **Responsibilization of patients** | Justice | Relates to patients adopting greater responsibility for their own health. Could also be fairness, but as justice also includes social justice this category was deemed to be a better fit. |
| **Responsibility** | Responsible Innovation | Synonym for/ dimension of existing theme |
| **Right of patient to know of algorithmic processing and to agree or object to such** | Consent | A nuance to or dimension of the existing theme. |
| **Risk of dehumanization** | Human Centeredness | |
| **Risk of inadequate governance in LMICs** | Distributive Justice | Relates to risk of exploitation / data colonialism by developers in the global north |
| **Risk of stigmatisation/ discrimination** | Fairness/bias | A nuance to or dimension of the existing theme. |

| | | |
|---|---|---|
| **Scientific integrity (honesty and rigour)** | Research Ethics | A nuance to or dimension of the existing theme. |
| **Shared responsibility for decision making** | Autonomy | Similar to 'human in the loop' arguments |
| **Standardisation (including quality standards)** | Accuracy | This is both positive- standards needed to secure accurate and appropriate outcomes and negative e.g. Standardisation via AI is also recognised as slipping from support to conditioning behaviours which is a problem for autonomy. However, in this context the discussion mainly focused on sense 1 |
| **Sustainability** | Sustainability | |
| **Traceability** | Transparency | A nuance to or dimension of the existing theme relating to data sources. |
| **Transparency of purpose for data collection for training and algorithm development** | Transparency | A nuance to or dimension of the existing theme relating to developers sourcing of data. |

| | | |
|---|---|---|
| **Trust** | Trust | Special cases and nuances listed across different papers: One argues that 'trust' in AI is not warranted and reliability is preferable. Another distinguishes rationally justified trust from value based trust. At least one paper discusses Trust more in relation to trust in companies' vs trust in regulations not AI itself. A paper on AI in Africa mentioned lack of trust of non-African developers using/stealing African data. |
| **Truthfulness** | Accuracy | Similar to loyalty, reliability etc- all relate to proper functioning of AI |
| **Validity** | Accuracy | Similar to Loyalty, reliability etc- all relate to proper functioning of AI |
| **Value of creating scientific knowledge as end in itself** | Human Centredness | |
| **Value of data (benefits of AI should benefit those patients who provided their data to build upon)** | Responsible Innovation / Distributive Justice | |
| **Veracity/ avoiding deception** | Accuracy | Similar to loyalty, reliability, truthfulness etc- all relate to proper functioning of AI |

**Table s6: Definitions of de novo ethical and social themes arising from coding of 'other' category in SR1**

| Ethical or societal theme | Consensus definition employed in charting SR1 | Notes |
|---|---|---|
| **Animal Justice** | Potential of AI to reduce animal suffering by replacing experimental animals with digital models | Only one instance in the whole data set but we considered it sufficiently novel to be worth highlighting |
| **Deskilling Risk** | Harm arising from loss of human capacities and practical knowledge as AI replaces human tasks | |
| **Dignity** | Cases where AI may specifically impact human dignity in ways that are not reducible to other existing themes in SR1. | |
| **Distributive Justice** | Subset of justice issues relating mainly to global access to AI and global distribution of benefits and harms of AI, especially with respect to the global north / global south | |
| **Human Centredness** | Need to orientate AI to human values and practices not the other way round. Incorporates threats to professional and personal identity, reduction of empathy and social cohesion due to automation of human interactions especially in the case of medical care | |

| | |
|---|---|
| **Sustainability** | Relates to both the economic and environmental impacts of AI. Systems that are not sustainable in both sense may be unethical as they can waste resources, lead patients and physicians to depend on them only to be discontinued and damage the environment |

**APPENDIX 6: REFERENCES FOR SR1 AND SR2**

**SR1**:
Althobaiti, K., & Althobaiti, K. (2021). Surveillance in Next-Generation Personalized Healthcare: Science and Ethics of Data Analytics in Healthcare. *NEW BIOETHICS-A MULTIDISCIPLINARY JOURNAL OF BIOTECHNOLOGY AND THE BODY*, *27*(4), 295-319. https://doi.org/10.1080/20502877.2021.1993055 EA OCT 2021

Arda, B. (2021). Ethico-legal aspects of artificial intelligence in neurocritical care). *European Journal of Neurology*, *28*, 27. https://doi.org/https://dx.doi.org/10.1111/ene.14970 PT - Conference Abstract (7th Congress of the European Academy of Neurology. Virtual.)

Arnold M.H. (2021). Teasing out Artificial Intelligence in Medicine: An Ethical Critique of Artificial Intelligence and Machine Learning in Medicine. *Journal of Bioethical Inquiry*, *18*(1), 121-139. https://doi.org/https://dx.doi.org/10.1007/s11673-020-10080-1 PT - Article

Babushkina, D., Votsis, A., Babushkina, D., & Votsis, A. (2022). Epistemo-ethical constraints on AI-human decision making for diagnostic purposes. *Ethics and Information Technology*, *24*(2). https://doi.org/10.1007/s10676-022-09629-y

Bach, S., Sezgin, E., Linwood, S. L., Bach, S., Sezgin, E., & Linwood, S. L. (2022). Ethical artificial intelligence in paediatrics. *LANCET CHILD & ADOLESCENT HEALTH*, *6*(12), 833-835. https://doi.org/10.1016/S2352-4642(22)00243-7 EA NOV 2022

Badal, K., Lee, C. M., Esserman, L. J., Badal, K., Lee, C. M., & Esserman, L. J. (2023). Guiding principles for the responsible development of artificial intelligence tools for healthcare. *COMMUNICATIONS MEDICINE*, *3*(1). https://doi.org/10.1038/s43856-023-00279-9

Baeroe, K., Gundersen, T., Henden, E., Rommetveit, K., Baeroe, K., Gundersen, T., Henden, E., & Rommetveit, K. (2022). Can medical algorithms be fair? Three ethical quandaries and one dilemma. *BMJ HEALTH & CARE INFORMATICS*, *29*(1). https://doi.org/10.1136/bmjhci-2021-100445

Bauer, G. R., & Lizotte, D. J. (2021). Artificial Intelligence, Intersectionality, and the Future of Public Health. *American journal of public health*, *111*(1), 98-100. https://doi.org/https://dx.doi.org/10.2105/AJPH.2020.306006 PT - Editorial

Bellini, V., Montomoli, J., Bignami E. (2021). Poor quality data, privacy, lack of certifications: the lethal triad of new technologies in intensive care. *Intensive Care Medicine*, *47*(9), 1052-1053. https://doi.org/https://dx.doi.org/10.1007/s00134-021-06473-4 PT - Letter

Bhattacharya, S., Hossain, M. M., Juyal, R., Sharma, N., Pradhan, K. B., Singh, A., Bhattacharya, S., Hossain, M. M., Juyal, R., Sharma, N., Pradhan, K. B., & Singh, A. (2021). Role of Public Health Ethics or Responsible Use of Artificial Intelligence Technologies. *INDIAN JOURNAL OF COMMUNITY MEDICINE*, *46*(2), 178-181. https://doi.org/10.4103/ijcm.IJCM_62_20

Biller-Andorno, N., Ferrario, A., Gloeckler, S., Biller-Andorno, N., Ferrario, A., & Gloeckler, S. (2022). In Search of a Mission: Artificial Intelligence in Clinical Ethics COMMENT. *American Journal of Bioethics*, *22*(7), 23-25. https://doi.org/10.1080/15265161.2022.2075055

Binkley, C. E., & Green, B. P. (2021). Does Intraoperative Artificial Intelligence Decision Support Pose Ethical Issues? *JAMA Surgery*, *156*(9), 809-810. https://doi.org/https://dx.doi.org/10.1001/jamasurg.2021.2055 PT - Note

Blumenthal-Barby, J., Lang, B., Dorfman, N., Kaplan, H., Hooper, W. B., Kostick-Quenet, K., Blumenthal-Barby, J., Lang, B., Dorfman, N., Kaplan, H., Hooper, W. B., & Kostick-Quenet, K. (2022). Research on the Clinical Translation of Health Care Machine Learning: Ethicists Experiences on Lessons Learned. *American Journal of Bioethics*, *22*(5), 1-3. https://doi.org/10.1080/15265161.2022.2059199

Bradley, E. A., Khan, A., McNeal, D. M., Bravo-Jaimes, K., Khanna, A., Cook, S., Opotowsky, A. R., John, A., Lee, M., Pasquali, S., Daniels, C. J., Pernick, M., Kirkpatrick, J. N., Gurvitz, M., Bradley, E. A., Khan, A., McNeal, D. M., Bravo-Jaimes, K., Khanna, A., . . . Gurvitz, M. (2022). Operational and Ethical Considerations for a National Adult Congenital Heart Disease Database. *JOURNAL OF THE AMERICAN HEART ASSOCIATION*, *11*(7). https://doi.org/10.1161/JAHA.121.022338

Braun, M., & Braun, M. (2022). Ethics of digital twins: four challenges. *Journal of Medical Ethics*, *48*(9), 579-580. https://doi.org/10.1136/medethics-2021-107675 EA AUG 2021

Braun, M., Hummel, P., Beck, S., Dabrock, P., Braun, M., Hummel, P., Beck, S., & Dabrock, P. (2021). Primer on an ethics of AI-based decision support systems in the clinic. *Journal of Medical Ethics*, *47*(12). https://doi.org/10.1136/medethics-2019-105860

Brown, J. E. H., Halpern J. (2021). AI chatbots cannot replace human interactions in the pursuit of more inclusive mental healthcare. *SSM - Mental Health*, *1*, 100017. https://doi.org/https://dx.doi.org/10.1016/j.ssmmh.2021.100017 PT - Article

Burns, T. F., & Burns, T. F. (2023). Epistemic virtues of harnessing rigorous machine learning systems in ethically sensitive domains. *Journal of Medical Ethics*. https://doi.org/10.1136/jme-2023-109105 EA MAY 2023

Capasso, M., Umbrello, S., Capasso, M., & Umbrello, S. (2022). Responsible nudging for social good: new healthcare skills for AI-driven digital personal assistants. *MEDICINE HEALTH CARE AND PHILOSOPHY*, *25*(1), 11-22. https://doi.org/10.1007/s11019-021-10062-z EA NOV 2021

Carruthers, R., Straw, I., Ruffle, J. K., Herron, D., Nelson, A., Bzdok, D., Fernandez-Reyes, D., Rees, G., Nachev, P., Carruthers, R., Straw, I., Ruffle, J. K., Herron, D., Nelson, A., Bzdok, D., Fernandez-Reyes, D., Rees, G., & Nachev, P. (2022). Representational ethical model calibration. *NPJ DIGITAL MEDICINE*, *5*(1). https://doi.org/10.1038/s41746-022-00716-4

Chan, B., & Chan, B. (2023). Black-box assisted medical decisions: AI power vs. ethical physician care. *MEDICINE HEALTH CARE AND PHILOSOPHY*. https://doi.org/10.1007/s11019-023-10153-z EA JUN 2023

Chen, I. Y., Pierson, E., Rose, S., Joshi, S., Ferryman, K., Ghassemi, M., Chen, I. Y., Pierson, E., Rose, S., Joshi, S., Ferryman, K., & Ghassemi, M. (2021). Ethical Machine Learning in Healthcare. ANNUAL REVIEW OF BIOMEDICAL DATA SCIENCE, VOL 4, 4139 EL CAMINO WAY, PO BOX 10139, PALO ALTO, CA 94303-0897 USA.

Chiang, S., Picard, R. W., Chiong, W., Moss, R., Worrell, G. A., Rao, V. R., Goldenholz D.M. (2021). Guidelines for Conducting Ethical Artificial Intelligence Research in Neurology: A Systematic Approach for Clinicians and Researchers. *Neurology*, *97*(13), 632-640. https://doi.org/https://dx.doi.org/10.1212/WNL.0000000000012570 PT - Article

Chiruvella, V., Guddati, A. K., Chiruvella, V., & Guddati, A. K. (2021). Ethical Issues in Patient Data Ownership. *INTERACTIVE JOURNAL OF MEDICAL RESEARCH*, *10*(2). https://doi.org/10.2196/22269

Cobianchi, L., Verde, J. M., Loftus, T. J., Piccolo, D., Dal Mas, F., Mascagni, P., Vazquez, A. G., Ansaloni, L., Marseglia, G. R., Massaro, M., Gallix, B., Padoy, N., Peter, A., Kaafarani, H. M., Cobianchi, L., Verde, J. M., Loftus, T. J., Piccolo, D., Dal Mas, F., . . . Kaafarani, H. M. (2022). Artificial Intelligence and Surgery: Ethical Dilemmas and Open Issues. *JOURNAL OF THE AMERICAN COLLEGE OF SURGEONS*, *235*(2), 268-275. https://doi.org/10.1097/XCS.0000000000000242

Coghlan, S., Leins, K., Sheldrick, S., Cheong, M., Gooding, P., D'Alfonso, S., Coghlan, S., Leins, K., Sheldrick, S., Cheong, M., Gooding, P., & D'Alfonso, S. (2023). To chat or bot to chat: Ethical issues with using chatbots in mental health. *DIGITAL HEALTH*, *9*. https://doi.org/10.1177/20552076231183542

Cordeiro, J. V. (2021). Digital Technologies and Data Science as Health Enablers: An Outline of Appealing Promises and Compelling Ethical, Legal, and Social Challenges. *FRONTIERS IN MEDICINE*, *8*, 647897. https://doi.org/https://dx.doi.org/10.3389/fmed.2021.647897 PT - Review

Cordero, D. A. C., & Cordero Jr, D. A. (2022). Artificial intelligence (AI) and human virtues: Towards a moral and quality health care system. *INTERNATIONAL JOURNAL FOR QUALITY IN HEALTH CARE*, *34*(4). https://doi.org/10.1093/intqhc/mzac096

Couture, V., Roy, M. C., Dez, E., Laperle, S., Belisle-Pipon, J. C., Couture, V., Roy, M.-C., Dez, E., Laperle, S., & Belisle-Pipon, J.-C. (2023). Ethical Implications of Artificial Intelligence in Population Health and the Public's Role in Its Governance: Perspectives From a Citizen and Expert Panel. *JOURNAL OF MEDICAL INTERNET RESEARCH*, *25*. https://doi.org/10.2196/44357

Currie, G., & Hawk, K. E. (2021). Ethical and Legal Challenges of Artificial Intelligence in Nuclear Medicine. *Seminars in Nuclear Medicine*, *51*(2), 120-125. https://doi.org/https://dx.doi.org/10.1053/j.semnuclmed.2020.08.001 PT - Review

Dave, T., Athaluri, S. A., Singh, S., Dave, T., Athaluri, S. A., & Singh, S. (2023). ChatGPT in medicine: an overview of its applications, advantages, limitations, future prospects, and ethical considerations. *FRONTIERS IN ARTIFICIAL INTELLIGENCE*, *6*. https://doi.org/10.3389/frai.2023.1169595

de Boer, B., & Kudina, O. (2021). What is morally at stake when using algorithms to make medical diagnoses? Expanding the discussion beyond risks and harms. *Theoretical Medicine and Bioethics*, *42*(5), 245-266.

De Manuel, A., Delgado, J., Iris, P. J., Ausín, T., Casacuberta, D., Cruz Piqueras, M., Guersenzvaig, A., Moyano, C., Rodríguez-Arias, D., Rueda, J., & Puyol, A. (2023). Ethical assessments and mitigation strategies for biases in AI-systems used during the COVID-19 pandemic. *Big Data and Society*, *10*(1).

De Simone, B., Di Saverio, S., De Simone, B., & Di Saverio, S. (2022). Artificial Intelligence in Surgical Care: We Must Overcome Ethical Boundaries. *JOURNAL OF THE AMERICAN COLLEGE OF SURGEONS*, *235*(2), 275-277. https://doi.org/10.1097/XCS.0000000000000227

Dhar, T., Dey, N., Borra, S., & Sherratt, R. S. (2023). Challenges of Deep Learning in Medical Image Analysis—Improving Explainability and Trust. *IEEE Transactions on Technology and Society*, *4*(1), 68-75. https://doi.org/10.1109/TTS.2023.3234203

Dlugatch, R., Georgieva, A., Kerasidou, A., Dlugatch, R., Georgieva, A., & Kerasidou, A. (2023). Trustworthy artificial intelligence and ethical design: public perceptions of trustworthiness of an AI-based decision-support tool in the context of intrapartum care. *BMC Medical Ethics*, *24*(1). https://doi.org/10.1186/s12910-023-00917-w

Donia, J., Shaw, J. A., Donia, J., & Shaw, J. A. (2021). Co-design and ethical artificial intelligence for health: An agenda for critical research and practice. *BIG DATA & SOCIETY*, *8*(2). https://doi.org/10.1177/20539517211065248

Dove, E. S., Reed-Berendt, R., Pareek, M., Dove, E., Reed-Berendt, R., Pareek, M., & Grp, U.-R. S. C. (2022). "Data makes the story come to life:" understanding the ethical and legal implications of Big Data research involving ethnic minority healthcare workers in the United Kingdom-a qualitative study. *BMC Medical Ethics*, *23*(1). https://doi.org/10.1186/s12910-022-00875-9

Dulhanty, A. (2021). Present value of future health data: Ethics of data collection and use. *Bulletin of the World Health Organization*, *99*(2), 162-163. https://doi.org/https://dx.doi.org/10.2471/BLT.19.237248 PT - Note

Duran, J. M., Jongsma K.R. (2021). Who is afraid of black box algorithms? On the epistemological and ethical basis of trust in medical AI. *Journal of Medical Ethics*. https://doi.org/https://dx.doi.org/10.1136/medethics-2020-106820
Esmonde, K., Roth, S., Walker, A., Esmonde, K., Roth, S., & Walker, A. (2023). A social and ethical framework for providing health information obtained from combining genetics and fitness tracking data. *TECHNOLOGY IN SOCIETY*, *74*. https://doi.org/10.1016/j.techsoc.2023.102297 EA JUN 2023

Felder, R. M. (2021). Coming to Terms with the Black Box Problem: How to Justify AI Systems in Health Care. *Hastings Center Report*, *51*(4), 38-45.

Ferrario, A., & Ferrario, A. (2022). Design publicity of black box algorithms: a support to the epistemic and ethical justifications of medical AI systems Comment. *Journal of Medical Ethics*, *48*(7), 492-494. https://doi.org/10.1136/medethics-2021-107482 EA MAY 2021

Ferrario, A., Gloeckler, S., Biller-Andorno, N., Ferrario, A., Gloeckler, S., & Biller-Andorno, N. (2023). Ethics of the algorithmic prediction of goal of care preferences: from theory to practice. *Journal of Medical Ethics*, *49*(3), 165-174. https://doi.org/10.1136/jme-2022-108371 EA NOV 2022

Fritzsche, M. C., Akyuz, K., Abadia, M. C., McLennan, S., Marttinen, P., Mayrhofer, M. T., Buyx, A. M., Fritzsche, M.-C., Akyuz, K., Cano Abadia, M., McLennan, S., Marttinen, P., Mayrhofer, M. T., & Buyx, A. M. (2023). Ethical layering in AI-driven polygenic risk scores-New complexities, new challenges. *FRONTIERS IN GENETICS*, *14*. https://doi.org/10.3389/fgene.2023.1098439

Funer, F., Liedtke, W., Tinnemeyer, S., Klausen, A. D., Schneider, D., Zacharias, H. U., Langanke, M., Salloch, S., Funer, F., Liedtke, W., Tinnemeyer, S., Klausen, A. D., Schneider, D., Zacharias, H. U., Langanke, M., & Salloch, S. (2023). Responsibility and decision-making authority in using clinical decision support systems: an empirical-ethical exploration of German prospective professionals' preferences and concerns. *Journal of Medical Ethics*. https://doi.org/10.1136/jme-2022-108814 EA MAY 2023

Giovanola, B., & Tiribelli, S. (2023). Beyond bias and discrimination: redefining the AI ethics principle of fairness in healthcare machine-learning algorithms. *AI and Society*, *38*(2), 549-563. https://www.ncbi.nlm.nih.gov/pmc/articles/PMC9123626/pdf/146_2022_Article_1455.pdf

Grote, T., & Grote, T. (2022). Randomised controlled trials in medical AI: ethical considerations. *Journal of Medical Ethics*, *48*(11), 899-906. https://doi.org/10.1136/medethics-2020-107166 EA MAY 2021

Grote, T., & Keeling, G. (2022). On algorithmic fairness in medical practice. *Cambridge Quarterly of Healthcare Ethics*, *31*(1), 83-94. https://www.cambridge.org/core/services/aop-cambridge-core/content/view/C961BFF6311D83FBEF851240EA56E9F0/S0963180121000839a.pdf/div-class-title-on-algorithmic-fairness-in-medical-practice-div.pdf

Gundersen, T., Baeroe, K., Gundersen, T., & Baeroe, K. (2022). The Future Ethics of Artificial Intelligence in Medicine: Making Sense of Collaborative Models. *Science and Engineering Ethics*, *28*(2). https://doi.org/10.1007/s11948-022-00369-2

Gupta, S., Kamboj, S., Bag, S., Gupta, S., Kamboj, S., & Bag, S. (2021). Role of Risks in the Development of Responsible Artificial Intelligence in the Digital Healthcare Domain. *INFORMATION SYSTEMS FRONTIERS*. https://doi.org/10.1007/s10796-021-10174-0 EA AUG 2021

Hantel, A., Clancy, D. D., Kehl, K. L., Marron, J. M., Van Allen, E. M., Abel, G. A., Hantel, A., Clancy, D. D., Kehl, K. L., Marron, J. M., Van Allen, E. M., & Abel, G. A. (2022). A Process Framework for Ethically Deploying Artificial Intelligence in Oncology. *JOURNAL OF CLINICAL ONCOLOGY*, *40*(34), 3907-+. https://doi.org/10.1200/JCO.22.01113

Harrer, S., & Harrer, S. (2023). Attention is not all you need: the complicated case of ethically using large language models in healthcare and medicine. *EBIOMEDICINE*, *90*. https://doi.org/10.1016/j.ebiom.2023.104512 EA MAR 2023

Hatherley, J., Sparrow, R., Hatherley, J., & Sparrow, R. (2022). Diachronic and synchronic variation in the performance of adaptive machine learning systems: the ethical challenges. *JOURNAL OF THE AMERICAN MEDICAL INFORMATICS ASSOCIATION*. https://doi.org/10.1093/jamia/ocac218 EA NOV 2022

Heyen, N. B., Salloch, S., Heyen, N. B., & Salloch, S. (2021). The ethics of machine learning-based clinical decision support: an analysis through the lens of professionalisation theory. *BMC Medical Ethics*, *22*(1). https://doi.org/10.1186/s12910-021-00679-3

Ho, C. W. L., Caals, K., Ho, C. W.-L., & Caals, K. (2021). A Call for an Ethics and Governance Action Plan to Harness the Power of Artificial Intelligence and Digitalization in Nephrology. *SEMINARS IN NEPHROLOGY*, *41*(3), 282-293. https://doi.org/10.1016/j.semnephrol.2021.05.009 EA JUL 2021

Ho, C. W. L., Malpani, R., Ho, C. W.-L., & Malpani, R. (2022). Scaling up the Research Ethics Framework for Healthcare Machine Learning as Global Health Ethics and Governance. *American Journal of Bioethics*, *22*(5), 36-38. https://doi.org/10.1080/15265161.2022.2055209

Huang, P. H., Kim, K. H., Schermer, M., Huang, P.-H., Kim, K.-H., & Schermer, M. (2022). Ethical Issues of Digital Twins for Personalized Health Care Service: Preliminary Mapping Study. *JOURNAL OF MEDICAL INTERNET RESEARCH*, *24*(1). https://doi.org/10.2196/33081

Iqbal, J. D., Krauthammer, M., Biller-Andorno, N., Iqbal, J. D., Krauthammer, M., & Biller-Andorno, N. (2022). The Use and Ethics of Digital Twins in Medicine. *JOURNAL OF LAW MEDICINE & ETHICS*, *50*(3), 583-596. https://doi.org/10.1017/jme.2022.97

Jackson, B. R., Ye, Y., Crawford, J. M., Becich, M. J., Roy, S., Botkin, J. R., de Baca, M. E., Pantanowitz, L., Jackson, B. R., Ye, Y., Crawford, J. M., Becich, M. J., Roy, S., Botkin, J. R., de Baca, M. E., & Pantanowitz, L. (2021). The Ethics of Artificial Intelligence in Pathology and Laboratory Medicine: Principles and Practice. *ACADEMIC PATHOLOGY*, *8*. https://doi.org/10.1177/2374289521990784

Johansson, J. V., Bentzen, H. B., Mascalzoni, D., Johansson, J. V., Bentzen, H. B., & Mascalzoni, D. (2022). What ethical approaches are used by scientists when sharing health data? An interview study. *BMC Medical Ethics*, *23*(1). https://doi.org/10.1186/s12910-022-00779-8

Kaur, A., Garg, R., & Gupta, P. (2021). Challenges facing AI and Big data for Resource-poor Healthcare System. 2021 Second International Conference on Electronics and Sustainable Communication Systems (ICESC),

Keeling, G., & Grote, T. (2022). Enabling Fairness in Healthcare Through Machine Learning. *Ethics and Information Technology*, *24*(3), 1-13.

Kellmeyer P. (2021). Artificial intelligence in basic and clinical neuroscience: Opportunities and ethical challenges. *Neuroforum*, *25*(4), 241-250. https://doi.org/https://dx.doi.org/10.1515/nf-2019-0018 PT - Review

Kempt, H., Nagel, S. K., Kempt, H., & Nagel, S. K. (2022). Responsibility, second opinions and peer-disagreement: ethical and epistemological challenges of using AI in clinical diagnostic contexts. *Journal of Medical Ethics*, *48*(4), 222-229. https://doi.org/10.1136/medethics-2021-107440 EA DEC 2021

Kenny, L. M., Nevin, M., Fitzpatrick K. (2021). Ethics and standards in the use of artificial intelligence in medicine on behalf of the Royal Australian and New Zealand College of Radiologists. *Journal of Medical Imaging and Radiation Oncology*, *65*(5), 486-494. https://doi.org/https://dx.doi.org/10.1111/1754-9485.13289 PT - Article

Kong, J. D., Akpudo, U. E., Effoduh, J. O., Bragazzi, N. L., Kong, J. D., Akpudo, U. E., Effoduh, J. O., & Bragazzi, N. L. (2023). Leveraging Responsible, Explainable, and Local Artificial Intelligence Solutions for Clinical Public Health in the Global South. *HEALTHCARE*, *11*(4). https://doi.org/10.3390/healthcare11040457

Kostick-Quenet, K. M., Cohen, I. G., Gerke, S., Lo, B., Antaki, J., Movahedi, F., Njah, H., Schoen, L., Estep, J. E., & Blumenthal-Barby, J. S. (2022). Mitigating Racial Bias in Machine Learning. *Journal of Law, Medicine and Ethics*, *50*(1), 92-100.

Kotsenas, A. L., Balthazar, P., Andrews, D., Geis, J. R., & Cook, T. S. (2021). Rethinking Patient Consent in the Era of Artificial Intelligence and Big Data. *Journal of the American College of Radiology*, *18*(1), 180-184. https://doi.org/https://dx.doi.org/10.1016/j.jacr.2020.09.022 PT - Note

Kudina, O., de Boer, B., Kudina, O., & de Boer, B. (2021). Co-designing diagnosis: Towards a responsible integration of Machine Learning decision-support systems in medical diagnostics. *JOURNAL OF EVALUATION IN CLINICAL PRACTICE*, *27*(3), 529-536. https://doi.org/10.1111/jep.13535 EA JAN 2021

Kuhler, M., & Kuehler, M. (2022). Exploring the phenomenon and ethical issues of AI paternalism in health apps. *Bioethics*, *36*(2), 194-200. https://doi.org/10.1111/bioe.12886 EA MAY 2021

Kundu, S. (2021). AI in medicine must be explainable. *NATURE MEDICINE*, *27*(8), 1328. https://doi.org/https://dx.doi.org/10.1038/s41591-021-01461-z PT - Article

Lang, M., Bernier, A., Knoppers, B. M., Lang, M., Bernier, A., & Knoppers, B. M. (2022). Artificial Intelligence in Cardiovascular Imaging: "Unexplainable" Legal and Ethical Challenges? *CANADIAN JOURNAL OF CARDIOLOGY*, *38*(2), 225-233. https://doi.org/10.1016/j.cjca.2021.10.009 EA JAN 2022

Lee, S. S. (2022). Philosophical evaluation of the conceptualisation of trust in the NHS' Code of Conduct for artificial intelligence-driven technology. *Journal of Medical Ethics*, *48*(4), 272-277.

Leimanis, A., Palkova, K., Leimanis, A., & Palkova, K. (2021). Ethical Guidelines for Artificial Intelligence in Healthcare from the Sustainable Development Perspective. *EUROPEAN JOURNAL OF SUSTAINABLE DEVELOPMENT*, *10*(1), 90-102. https://doi.org/10.14207/ejsd.2021.v10n1p90

Levi, M., Bernstein, M., Waeiss, C., Levi, M., Bernstein, M., & Waeiss, C. (2022). Broadening the Ethical Scope. *American Journal of Bioethics*, *22*(5), 26-28. https://doi.org/10.1080/15265161.2022.2055219

Liao, S. M., & Liao, S. M. (2023). Ethics of AI and Health Care: Towards a Substantive Human Rights Framework. *TOPOI-AN INTERNATIONAL REVIEW OF PHILOSOPHY*, *42*(3), 857-866. https://doi.org/10.1007/s11245-023-09911-8 EA APR 2023

Liu, T. Y. A., Wu, J. H., Liu, T. Y. A., & Wu, J.-H. (2022). The Ethical and Societal Considerations for the Rise of Artificial Intelligence and Big Data in Ophthalmology. *FRONTIERS IN MEDICINE*, *9*. https://doi.org/10.3389/fmed.2022.845522

Lucivero, F., Hallowell, N., Lucivero, F., & Hallowell, N. (2021). Digital/computational phenotyping: What are the differences in the science and the ethics? *BIG DATA & SOCIETY*, *8*(2). https://doi.org/10.1177/20539517211062885

Makridis, C., Hurley, S., Klote, M., Alterovitz, G., Makridis, C., Hurley, S., Klote, M., & Alterovitz, G. (2021). Ethical Applications of Artificial Intelligence: Evidence From Health Research on Veterans. *JMIR MEDICAL INFORMATICS*, *9*(6). https://doi.org/10.2196/28921

Martinez-Martin, N., Greely, H. T., Cho, M. K., Martinez-Martin, N., Greely, H. T., & Cho, M. K. (2021). Ethical Development of Digital Phenotyping Tools for Mental Health Applications: Delphi Study. *JMIR MHEALTH AND UHEALTH*, *9*(7). https://doi.org/10.2196/27343

Martinez-Martin, N., Luo, Z., Kaushal, A., Adeli, E., Haque, A., Kelly, S. S., Wieten, S., Cho, M. K., Magnus, D., Fei-Fei, L., Schulman, K., & Milstein, A. (2021). Ethical issues in using ambient intelligence in health-care settings. *The Lancet Digital Health*, *3*(2), e115-e123. https://doi.org/https://dx.doi.org/10.1016/S2589-7500%2820%2930275-2 PT - Review

Martinho, A., Kroesen, M., & Chorus, C. (2021). A healthy debate: Exploring the views of medical doctors on the ethics of artificial intelligence. *Artificial Intelligence in Medicine*, *121*, 102190. https://doi.org/https://dx.doi.org/10.1016/j.artmed.2021.102190 PT - Article

McCoy, M. S., Allen, A. L., Kopp, K., Mello, M. M., Patil, D. J., Ossorio, P., Joffe, S., Emanuel, E. J., (2023). Ethical Responsibilities for Companies That Process Personal Data. *American Journal of Bioethics*. https://doi.org/10.1080/15265161.2023.2209535 EA MAY 2023

McCradden, M., Hui, K., Buchman, D. Z., McCradden, M., Hui, K., & Buchman, D. Z. (2022). Evidence, ethics and the promise of artificial intelligence in psychiatry. *Journal of Medical Ethics*. https://doi.org/10.1136/jme-2022-108447 EA DEC 2022

McCradden, M. D., Anderson, J. A., Stephenson, E. A., Drysdale, E., Erdman, L., Goldenberg, A., Shaul, R. Z., McCradden, M. D., Anderson, J. A., Stephenson, E. A., Drysdale, E., Erdman, L., Goldenberg, A., & Shaul, R. Z. (2022). A Research Ethics Framework for the Clinical Translation of Healthcare Machine Learning. *American Journal of Bioethics*, *22*(5), 8-22. https://doi.org/10.1080/15265161.2021.2013977 EA JAN 2022

McKay, F., Williams, B. J., Prestwich, G., Bansal, D., Hallowell, N., Treanor, D., McKay, F., Williams, B. J., Prestwich, G., Bansal, D., Hallowell, N., & Treanor, D. (2022). The ethical challenges of artificial intelligence-driven digital pathology. *JOURNAL OF PATHOLOGY CLINICAL RESEARCH*, *8*(3), 209-216. https://doi.org/10.1002/cjp2.263 EA FEB 2022

McKay, F., Williams, B. J., Prestwich, G., Bansal, D., Treanor, D., Hallowell, N., McKay, F., Williams, B. J., Prestwich, G., Bansal, D., Treanor, D., & Hallowell, N. (2023). Artificial intelligence and medical research databases: ethical review by data access committees. *BMC Medical Ethics*, *24*(1). https://doi.org/10.1186/s12910-023-00927-8

McKeown, A., Mourby, M., Harrison, P., Walker, S., Sheehan, M., Singh, I., McKeown, A., Mourby, M., Harrison, P., Walker, S., Sheehan, M., & Singh, I. (2021). Ethical Issues in Consent for the Reuse of Data in Health Data Platforms. *Science and Engineering Ethics*, *27*(1). https://doi.org/10.1007/s11948-021-00282-0

McLennan, S., Fiske, A., Tigard, D., Muller, R., Haddadin, S., Buyx, A., McLennan, S., Fiske, A., Tigard, D., Muller, R., Haddadin, S., & Buyx, A. (2022). Embedded ethics: a proposal for integrating ethics into the development of medical AI. *BMC Medical Ethics*, *23*(1). https://doi.org/10.1186/s12910-022-00746-3

Michelson, K. N., Klugman, C. M., Kho, A. N., Gerke, S., Michelson, K. N., Klugman, C. M., Kho, A. N., & Gerke, S. (2022). Ethical Considerations Related to Using Machine Learning-Based Prediction of Mortality in the Pediatric Intensive Care Unit. *JOURNAL OF PEDIATRICS*, *247*, 125-128. https://doi.org/10.1016/j.jpeds.2021.12.069

Miller, M., & Miller, M., Jr. (2022). Catholic Health Care and AI Ethics: Algorithms for Human Flourishing. *LINACRE QUARTERLY*, *89*(2), 152-164. https://doi.org/10.1177/00243639221082226

Milosevic, Z. (2021). Enabling scalable AI for Digital Health: interoperability, consent and ethics support. 2021 IEEE 25th International Enterprise Distributed Object Computing Workshop (EDOCW),

Milton C.L. (2021). Risking Human Dignity With Innovations: Artificial Intelligence and the Future of the Discipline of Nursing. *Nursing science quarterly*, *34*(3), 244-246. https://doi.org/https://dx.doi.org/10.1177/08943184211010440 PT - Article

Molldrem, S., Smith, A. K. J., McClelland, A., Molldrem, S., Smith, A. K. J., & McClelland, A. (2023). Predictive analytics in HIV surveillance require new approaches to data ethics, rights, and regulation in public health. *CRITICAL PUBLIC HEALTH*, *33*(3), 275-281. https://doi.org/10.1080/09581596.2022.2113035 EA AUG 2022

Morgan, M. B., Mates, J. L., Morgan, M. B., & Mates, J. L. (2023). Ethics of Artificial Intelligence in Breast Imaging. *JOURNAL OF BREAST IMAGING*, *5*(2), 195-200. https://doi.org/10.1093/jbi/wbac076 EA JAN 2023

Morris, M. X., Song, E. Y., Rajesh, A., Asaad, M., Phillips, B. T., Morris, M. X., Song, E. Y., Rajesh, A., Asaad, M., & Phillips, B. T. (2023). Ethical, Legal, and Financial Considerations of Artificial Intelligence in Surgery. *AMERICAN SURGEON*, *89*(1), 55-60. https://doi.org/10.1177/00031348221117042 EA AUG 2022

Mouchabac, S., Adrien, V., Falala-Sechet, C., Bonnot, O., Maatoug, R., Millet, B., Peretti, C. S., Bourla, A., Ferreri, F., Mouchabac, S., Adrien, V., Falala-Sechet, C., Bonnot, O., Maatoug, R., Millet, B., Peretti, C.-S., Bourla, A., & Ferreri, F. (2021). Psychiatric Advance Directives and Artificial Intelligence: A Conceptual Framework for Theoretical and Ethical Principles. *FRONTIERS IN PSYCHIATRY*, *11*. https://doi.org/10.3389/fpsyt.2020.622506

Muller, H., Mayrhofer, M. T., Evert-Ben Van, V., Holzinger, A., Mueller, H., Mayrhofer, M. T., Evert-Ben Van, V., & Holzinger, A. (2021). The Ten Commandments of Ethical Medical AI. *COMPUTER*, *54*(7), 119-123. https://doi.org/10.1109/MC.2021.3074263

Mykhailov, D.(2021). A moral analysis of intelligent decision-support systems in diagnostics through the lens of Luciano Floridi's information ethics. *HUMAN AFFAIRS-POSTDISCIPLINARY HUMANITIES & SOCIAL SCIENCES QUARTERLY*, *31*(2), 149-164. https://doi.org/10.1515/humaff-2021-0013

Naik, N., Hameed, B. M. Z., Shetty, D. K., Swain, D., Shah, M. L., Paul, R., Aggarwal, K., Ibrahim, S., Patil, V., Smriti, K., Shetty, S., Rai, B. P., Chlosta, P., Somani, B. K., Naik, N., Hameed, B. M. Z., Shetty, D. K., Swain, D., Shah, M., . . . Somani, B. K. (2022). Legal and Ethical Consideration in Artificial Intelligence in Healthcare: Who Takes Responsibility? *FRONTIERS IN SURGERY*, *9*. https://doi.org/10.3389/fsurg.2022.862322

Ng, M. Y., Kapur, S., Blizinsky, K. D., Hernandez-Boussard, T., Ng, M. Y., Kapur, S., Blizinsky, K. D., & Hernandez-Boussard, T. (2022). The AI life cycle: a holistic approach to creating ethical AI for health decisions Comment. *NATURE MEDICINE*, *28*(11), 2247-2249. https://doi.org/10.1038/s41591-022-01993-y EA SEP 2022

Nichol, A. A., Bendavid, E., Mutenherwa, F., Patel, C., Cho, M. K., Nichol, A. A., Bendavid, E., Mutenherwa, F., Patel, C., & Cho, M. K. (2021). Diverse experts' perspectives on ethical issues of using machine learning to predict HIV/AIDS risk in sub-Saharan Africa: a modified Delphi study. *BMJ OPEN*, *11*(7). https://doi.org/10.1136/bmjopen-2021-052287

Obasa, A. E., Palk, A. C., Obasa, A. E., & Palk, A. C. (2023). Responsible application of artificial intelligence in health care. *SOUTH AFRICAN JOURNAL OF SCIENCE*, *119*(5). https://doi.org/10.17159/sajs.2023/14889

Olczak, J., Pavlopoulos, J., Prijs, J., Ijpma, F. F. A., Doornberg, J. N., Lundstrom, C., Hedlund, J., & Gordon, M. (2021). Presenting artificial intelligence, deep learning, and machine learning studies to clinicians and healthcare stakeholders: an introductory reference with a guideline and a Clinical AI Research (CAIR) checklist proposal. *Acta Orthopaedica*, *92*(5), 513-525. https://doi.org/https://dx.doi.org/10.1080/17453674.2021.1918389 PT - Article

Oprescu, A. M., Miro-Amarante, G., Garcia-Diaz, L., Rey, V. E., Chimenea-Toscano, A., Martinez-Martinez, R., Romero-Ternero, M. C., Oprescu, A. M., Miro-Amarante, G., Garcia-Diaz, L., Rey, V. E., Chimenea-Toscano, A., Martinez-Martinez, R., & Romero-Ternero, M. C. (2022). Towards a data collection methodology for Responsible Artificial Intelligence in health: A prospective and qualitative study in pregnancy. *INFORMATION FUSION*, *83*, 53-78. https://doi.org/10.1016/j.inffus.2022.03.011 EA APR 2022

Ozdemir, V. (2021). Digital Is Political: Why We Need a Feminist Conceptual Lens on Determinants of Digital Health. *OMICS A Journal of Integrative Biology*, *25*(4), 249-254. https://doi.org/https://dx.doi.org/10.1089/omi.2021.0020 PT - Article

Padmapriya, S. T., Parthasarathy, S., Padmapriya, S. T., & Parthasarathy, S. (2023). Ethical Data Collection for Medical Image Analysis: a Structured Approach. *Asian Bioethics Review*. https://doi.org/10.1007/s41649-023-00250-9 EA APR 2023

Palmer, A., Schwan, D., Palmer, A., & Schwan, D. (2023). More Process, Less Principles: The Ethics of Deploying AI and Robotics in Medicine. *Cambridge Quarterly of Healthcare Ethics*. https://doi.org/10.1017/S0963180123000087 EA APR 2023

Parviainen, J., Rantala, J., Parviainen, J., & Rantala, J. (2022). Chatbot breakthrough in the 2020s? An ethical reflection on the trend of automated consultations in health care. *MEDICINE HEALTH CARE AND PHILOSOPHY*, *25*(1), 61-71. https://doi.org/10.1007/s11019-021-10049-w EA SEP 2021

Pennestri, F., Banfi, G., Pennestri, F., & Banfi, G. (2022). Artificial intelligence in laboratory medicine: fundamental ethical issues and normative key-points. *CLINICAL CHEMISTRY AND LABORATORY MEDICINE*, *60*(12), 1867-1874. https://doi.org/10.1515/cclm-2022-0096 EA APR 2022

Polyakova, I., & Sysoev, A. (2023). Ethical Problems of Applying Artificial Intelligence: Medical Intelligent Systems and Autonomous Vehicles. 2023 3rd International Conference on Technology Enhanced Learning in Higher Education (TELE),

Popa, E. O., van Hilten, M., Oosterkamp, E., Bogaardt, M. J., Popa, E. O., van Hilten, M., Oosterkamp, E., & Bogaardt, M.-J. (2021). The use of digital twins in healthcare: socio-ethical benefits and socio-ethical risks. *LIFE SCIENCES SOCIETY AND POLICY*, *17*(1). https://doi.org/10.1186/s40504-021-00113-x

Pourzolfaghar, Z., Alfano, M., Helfert, M., Pourzolfaghar, Z., Alfano, M., & Helfert, M. (2023). Application of ethical AI requirements to an AI solution use-case in healthcare domain. *AMERICAN JOURNAL OF BUSINESS*. https://doi.org/10.1108/AJB-12-2022-0201 EA JUN 2023

Predel, C., & Steger, F. (2021). Ethical Challenges With Smartwatch-Based Screening for Atrial Fibrillation: Putting Users at Risk for Marketing Purposes? *FRONTIERS IN CARDIOVASCULAR MEDICINE*, *7*, 615927. https://doi.org/https://dx.doi.org/10.3389/fcvm.2020.615927 PT - Article

Rathkopf, C., Heinrichs, B., Rathkopf, C., & Heinrichs, B. (2023). Learning to Live with Strange Error: Beyond Trustworthiness in Artificial Intelligence Ethics. *Cambridge Quarterly of Healthcare Ethics*. https://doi.org/10.1017/S0963180122000688 EA JAN 2023

Rauter, C. M., Wohlke, S., Schicktanz, S., Rauter, C. M., Woehlke, S., & Schicktanz, S. (2021). My Data, My Choice? - German Patient Organizations' Attitudes towards Big Data-Driven Approaches in Personalized Medicine. An Empirical-Ethical Study. *JOURNAL OF MEDICAL SYSTEMS*, *45*(4). https://doi.org/10.1007/s10916-020-01702-7

Richardson, J. P., Smith, C., Curtis, S., Watson, S., Zhu, X., Barry, B., Sharp R.R. (2021). Patient apprehensions about the use of artificial intelligence in healthcare. *NPJ DIGITAL MEDICINE*, *4*(1), 140. https://doi.org/https://dx.doi.org/10.1038/s41746-021-00509-1 PT - Article

Ronquillo, C. E., Peltonen, L. M., Pruinelli, L., Chu, C. H., Bakken, S., Beduschi, A., Cato, K., Hardiker, N., Junger, A., Michalowski, M., Nyrup, R., Rahimi, S., Reed, D. N., Salakoski, T., Salantera, S., Walton, N., Weber, P., Wiegand, T., Topaz M. (2021). Artificial intelligence in nursing: Priorities and opportunities from an international invitational think-tank of the Nursing and Artificial Intelligence Leadership Collaborative. *Journal of advanced nursing*, *77*(9), 3707-3717. https://doi.org/https://dx.doi.org/10.1111/jan.14855 PT - Article

Rubeis, G., Fang, M. L., & Sixsmith, A. (2022). Equity in AgeTech for Ageing Well in Technology-Driven Places: The Role of Social Determinants in Designing AI-based Assistive Technologies. *Science and Engineering Ethics*, *28*(6), 1-15.

Rubeis, G., & Rubeis, G. (2022). iHealth: The ethics of artificial intelligence and big data in mental healthcare. *INTERNET INTERVENTIONS-THE APPLICATION OF INFORMATION TECHNOLOGY IN MENTAL AND BEHAVIOURAL HEALTH*, *28*. https://doi.org/10.1016/j.invent.2022.100518 EA MAR 2022

Rueda, J., Delgado Rodríguez, J., Parra Jounou, I., Hortal-Carmona, J., Ausín, T., & Rodríguez-Arias, D. (2022). "Just" accuracy? Procedural fairness demands explainability in AI-based medical resource allocation. *AI and Society*.

Sabatello, M., & Sabatello, M. (2022). Wrongful Birth: AI-Tools for Moral Decisions in Clinical Care in the Absence of Disability Ethics. *American Journal of Bioethics*, *22*(7), 43-46. https://doi.org/10.1080/15265161.2022.2075971

Samuel, G., Chubb, J., Derrick G. (2021). Boundaries Between Research Ethics and Ethical Research Use in Artificial Intelligence Health Research. *Journal of empirical research on human research ethics : JERHRE*, *16*(3), 325-337. https://doi.org/https://dx.doi.org/10.1177/15562646211002744 PT - Article

Sand, M., Duran, J. M., Jongsma, K. R., Sand, M., Duran, J. M., & Jongsma, K. R. (2022). Responsibility beyond design: Physicians' requirements for ethical medical AI. *Bioethics*, *36*(2), 162-169. https://doi.org/10.1111/bioe.12887 EA JUN 2021

Scherz, P., & Scherz, P. (2022). Data Ethics, AI, and Accompaniment: The Dangers of Depersonalization in Catholic Health Care. *THEOLOGICAL STUDIES*, *83*(2), 271-292. https://doi.org/10.1177/00405639221096770 EA JUN 2022

Shaw, J., & Shaw, J. (2022). Emerging Paradigms for Ethical Review of Research Using Artificial Intelligence. *American Journal of Bioethics*, *22*(5), 42-44. https://doi.org/10.1080/15265161.2022.2055206

Shaw, J. A., Sethi, N., Block B.L. (2021). Five things every clinician should know about AI ethics in intensive care. *Intensive Care Medicine*, *47*(2), 157-159. https://doi.org/https://dx.doi.org/10.1007/s00134-020-06277-y PT - Editorial

Shen, F. X., Silverman, B. C., Monette, P., Kimble, S., Rauch, S. L., Baker, J. T., Shen, F. X., Silverman, B. C., Monette, P., Kimble, S., Rauch, S. L., & Baker, J. T. (2022). An Ethics Checklist for Digital Health Research in Psychiatry: Viewpoint. *JOURNAL OF MEDICAL INTERNET RESEARCH*, *24*(2). https://doi.org/10.2196/31146

Sidebottom, R., Lyburn, I., Brady, M., Vinnicombe S. (2021). Fair shares: building and benefiting from healthcare AI with mutually beneficial structures and development partnerships. *British Journal of Cancer*, *125*(9), 1181-1184. https://doi.org/https://dx.doi.org/10.1038/s41416-021-01454-2 PT - Review

Simon, D. A., Evans, B. J., Shachar, C., Cohen, G., Simon, D. A., Evans, B. J., Shachar, C., & Cohen, G. (2022). Should Alexa diagnose Alzheimer's?: Legal and ethical issues with at-home consumer devices. *CELL REPORTS MEDICINE*, *3*(12). https://doi.org/10.1016/j.xcrm.2022.100692 EA DEC 2022

Spector-Bagdady, K., Rahimzadeh, V., Jaffe, K., Moreno, J., Spector-Bagdady, K., Rahimzadeh, V., Jaffe, K., & Moreno, J. (2022). Promoting Ethical Deployment of Artificial Intelligence and Machine Learning in Healthcare. *American Journal of Bioethics*, *22*(5), 4-7. https://doi.org/10.1080/15265161.2022.2059206

Spiegel, J. M., Wilkinson, J., Ehrlich, R., Lockhart, K., Yassi, A., Barker, S., Riera, F., Kistnasamy B. (2021). Using artificial intelligence for high-volume identification of silicosis and tuberculosis a bio-ethics approach. *Annals of Global Health*, *87*(1), 58. https://doi.org/https://dx.doi.org/10.5334/aogh.3206 PT - Article

Sqalli, M. T., Aslonov, B., Gafurov, M., Nurmatov, S., Sqalli, M. T., Aslonov, B., Gafurov, M., & Nurmatov, S. (2023). Humanizing AI in medical training: ethical framework for responsible design. *FRONTIERS IN ARTIFICIAL INTELLIGENCE*, *6*. https://doi.org/10.3389/frai.2023.1189914

Stake, M., Heinrichs, B., Stake, M., & Heinrichs, B. (2022). Ethical Implications of e-Health Applications in Early Preventive Healthcare. *FRONTIERS IN GENETICS*, *13*. https://doi.org/10.3389/fgene.2022.902631

Starke, G., De Clercq, E., Borgwardt, S., Elger, B. S., Starke, G., De Clercq, E., Borgwardt, S., & Elger, B. S. (2021). Computing schizophrenia: ethical challenges for machine learning in psychiatry. *PSYCHOLOGICAL MEDICINE*, *51*(15), 2515-2521. https://doi.org/10.1017/S0033291720001683

Starke, G., De Clercq, E., & Elger, B. S. (2021). Towards a pragmatist dealing with algorithmic bias in medical machine learning. *Medicine, Health Care and Philosophy*, *24*(3), 341-349.

Stewart, C., Wong, S. K. Y., Sung J.J.Y. (2021). Mapping ethico-legal principles for the use of artificial intelligence in gastroenterology. *Journal of Gastroenterology and Hepatology (Australia)*, *36*(5), 1143-1148. https://doi.org/https://dx.doi.org/10.1111/jgh.15521 PT - Review

Svensson, A. M., & Jotterand, F. (2022). Doctor Ex Machina: A Critical Assessment of the Use of Artificial Intelligence in Health Care. *Journal of Medicine and Philosophy*, *47*(1), 155-178.

Szabo, L., Raisi-Estabragh, Z., Salih, A., McCracken, C., Pujadas, E. R., Gkontra, P., Kiss, M., Maurovich-Horvath, P., Vago, H., Merkely, B., Lee, A. M., Lekadir, K., Petersen, S. E., Szabo, L., Raisi-Estabragh, Z., Salih, A., McCracken, C., Pujadas, E. R., Gkontra, P., . . . Petersen, S. E. (2022). Clinician's guide to trustworthy and responsible artificial intelligence in cardiovascular imaging. *FRONTIERS IN CARDIOVASCULAR MEDICINE*, *9*. https://doi.org/10.3389/fcvm.2022.1016032

Thomasian, N. M., Eickhoff, C., Adashi E.Y. (2021). Advancing health equity with artificial intelligence. *Journal of public health policy*, *42*(4), 602-611. https://doi.org/https://dx.doi.org/10.1057/s41271-021-00319-5 PT - Review

Ti, L., Ho, A., & Knight, R. (2021). Towards Equitable AI Interventions for People Who Use Drugs: Key Areas That Require Ethical Investment. *Journal of Addiction Medicine*, *15*(2), 96-98. https://doi.org/https://dx.doi.org/10.1097/ADM.0000000000000722 PT - Review

Uusitalo, S., Tuominen, J., Arstila V. Ao. (2021). Mapping out the philosophical questions of AI and clinical practice in diagnosing and treating mental disorders. *JOURNAL OF EVALUATION IN CLINICAL PRACTICE*, *27*(3), 478-484. https://doi.org/https://dx.doi.org/10.1111/jep.13485 PT - Conference Paper

van der Niet, A. G., Bleakley A. (2021). Where medical education meets artificial intelligence: 'Does technology care?'. *Medical education*, *55*(1), 30-36. https://doi.org/https://dx.doi.org/10.1111/medu.14131 PT - Article

Van Der Veer, S. N., Riste, L., Cheraghi-Sohi, S., Phipps, D. L., Tully, M. P., Bozentko, K., Atwood, S., Hubbard, A., Wiper, C., Oswald, M., Peek N. Ao. (2021). Trading off accuracy and explainability in AI decision-making: findings from 2 citizens' juries. *Journal of the American Medical Informatics Association*, *28*(10), 2128-2138. https://doi.org/https://dx.doi.org/10.1093/jamia/ocab127 PT - Article

van Grondelle, J., Timmers, C., van Gorp, A., van Steenbergen, M., & Strikwerda, L. (2022). The value sensitive design of a preventive health check app. *Ethics and Information Technology*, *24*(3), 1-12.

Vandemeulebroucke, T., Denier, Y., Gastmans, C., Vandemeulebroucke, T., Denier, Y., & Gastmans, C. (2022). The Need for a Global Approach to the Ethical Evaluation of Healthcare Machine Learning. *American Journal of Bioethics*, *22*(5), 33-35. https://doi.org/10.1080/15265161.2022.2055207

Venkatachalam, S., Padmavathi, T., Vinodh, N., Thilagavathi, J., Joshi, G., Ramachandran, G., & Rajasekaran, B. (2023). Analysis of Ethical Issues Associated with Wearable Medical Devices. 2023 5th International Conference on Smart Systems and Inventive Technology (ICSSIT),

Vilaza, G. N., & McCashin, D. (2021). Is the Automation of Digital Mental Health Ethical? Applying an Ethical Framework to Chatbots for Cognitive Behaviour Therapy. *Frontiers in Digital Health*, *3*, 689736. https://doi.org/https://dx.doi.org/10.3389/fdgth.2021.689736 PT - Article

Wadden J.J. (2021). Defining the undefinable: the black box problem in healthcare artificial intelligence. *Journal of Medical Ethics*. https://doi.org/https://dx.doi.org/10.1136/medethics-2021-107529 PT - Article

Wang, W. S., Chen, L., Xiong, M. R., Wang, Y. C., Wang, W., Chen, L., Xiong, M., & Wang, Y. (2021). Accelerating AI Adoption with Responsible AI Signals and Employee Engagement Mechanisms in Health Care. *INFORMATION SYSTEMS FRONTIERS*. https://doi.org/10.1007/s10796-021-10154-4 EA JUN 2021

Weissglass, D. (2022). Contextual bias, the democratization of healthcare, and medical artificial intelligence in low- and middle-income countries. *Bioethics*.

Willem, T., Krammer, S., Bohm, A. S., French, L. E., Hartmann, D., Lasser, T., Buyx, A. (2022). Risks and benefits of dermatological machine learning health care applications-an overview and ethical analysis. *JOURNAL OF THE EUROPEAN ACADEMY OF DERMATOLOGY AND VENEREOLOGY*, *36*(9), 1660-1668. https://doi.org/10.1111/jdv.18192 EA MAY 2022

Yang, C. (2022). Digital contact tracing in the pandemic cities: Problematizing the regime of traceability in South Korea. *Big Data and Society*, *9*(1).

Youssef, A., Abramoff, M., Char, D., Youssef, A., Abramoff, M., & Char, D. (2023). Is the Algorithm Good in a Bad World, or Has It Learned to be Bad? The Ethical Challenges of "Locked" Versus "Continuously Learning" and "Autonomous" Versus "Assistive" AI Tools in Healthcare. *American Journal of Bioethics*, *23*(5), 43-45. https://doi.org/10.1080/15265161.2023.2191052

Zarif, A.(2022). The ethical challenges facing the widespread adoption of digital healthcare technology. *HEALTH AND TECHNOLOGY*, *12*(1), 175-179. https://doi.org/10.1007/s12553-021-00596-w EA OCT 2021

Zhang, J., Zhang, Z. M. (2023). Ethics and governance of trustworthy medical artificial intelligence. *BMC MEDICAL INFORMATICS AND DECISION MAKING*, *23*(1). https://doi.org/10.1186/s12911-023-02103-9

**SR2**:

Al-Hwsali, A., Alsaadi, B., Abdi, N., Khatab, S., Alzubaidi, M., Solaiman, B., & Househ, M. Scoping Review: Legal and Ethical Principles of Artificial Intelligence in Public Health. *Studies in Health Technology & Informatics*, *305*, 640-643. https://doi.org/https://dx.doi.org/10.3233/SHTI230579

Benzinger, L., Ursin, F., Balke, W. T., Kacprowski, T., Salloch, S., Benzinger, L., Ursin, F., Balke, W.-T., Kacprowski, T., & Salloch, S. (2023). Should Artificial Intelligence be used to support clinical ethical decision-making? A systematic review of reasons. *BMC Medical Ethics*, *24*(1). https://doi.org/10.1186/s12910-023-00929-6

Cartolovni, A., Tomicic, A., Mosler, E. L., Cartolovni, A., Tomicic, A., & Mosler, E. L. (2022). Ethical, legal, and social considerations of AI-based medical decision-support tools: A scoping review. *International Journal of Medical Informatics*, *161*. https://doi.org/10.1016/j.ijmedinf.2022.104738 EA MAR 2022

d'Elia, A., Gabbay, M., Rodgers, S., Kierans, C., Jones, E., Durrani, I., Thomas, A., Frith, L., d'Elia, A., Gabbay, M., Rodgers, S., Kierans, C., Jones, E., Durrani, I., Thomas, A., & Frith, L. (2022). Artificial intelligence and health inequities in primary care: a systematic scoping review and framework. *FAMILY MEDICINE AND COMMUNITY HEALTH*, *10*. https://doi.org/10.1136/fmch-2022-001670

Favaretto, M., Shaw, D., De Clercq, E., Joda, T., Elger, B. S., Favaretto, M., Shaw, D., De Clercq, E., Joda, T., & Elger, B. S. (2020). Big Data and Digitalization in Dentistry: A Systematic Review of the Ethical Issues. *INTERNATIONAL JOURNAL OF ENVIRONMENTAL RESEARCH AND PUBLIC HEALTH*, *17*(7). https://doi.org/10.3390/ijerph17072495

Ford, E., Shepherd, S., Jones, K., Hassan, L., Ford, E., Shepherd, S., Jones, K., & Hassan, L. (2021). Toward an Ethical Framework for the Text Mining of Social Media for Health Research: A Systematic Review. *Frontiers in Digital Health*, *2*. https://doi.org/10.3389/fdgth.2020.592237

Goirand, M., Austin, E., Clay-Williams, R., Goirand, M., Austin, E., & Clay-Williams, R. (2021). Implementing Ethics in Healthcare AI-Based Applications: A Scoping Review. *Science and Engineering Ethics*, *27*(5). https://doi.org/10.1007/s11948-021-00336-3

Goisauf, M., & Cano Abadia, M. Ethics of AI in Radiology: A Review of Ethical and Societal Implications. *Frontiers in Big Data*, *5*, 850383. https://doi.org/https://dx.doi.org/10.3389/fdata.2022.850383

Gooding, P., Kariotis, T., Gooding, P., & Kariotis, T. (2021). Ethics and Law in Research on Algorithmic and Data-Driven Technology in Mental Health Care: Scoping Review. *JMIR Mental Health*, *8*(6). https://doi.org/10.2196/24668

Ienca, M., Ferretti, A., Hurst, S., Puhan, M., Lovis, C., Vayena, E., Ienca, M., Ferretti, A., Hurst, S., Puhan, M., Lovis, C., & Vayena, E. (2018). Considerations for ethics review of big data health research: A scoping review. *PLOS ONE*, *13*(10). https://doi.org/10.1371/journal.pone.0204937

Istasy, P., Lee, W. S., Iansavichene, A., Upshur, R., Gyawali, B., Burkell, J., Sadikovic, B., Lazo-Langner, A., Chin-Yee, B., Istasy, P., Lee, W. S., Iansavichene, A., Upshur, R., Gyawali, B., Burkell, J., Sadikovic, B., Lazo-Langner, A., & Chin-Yee, B. (2022). The Impact of Artificial Intelligence on Health Equity in Oncology: Scoping Review. *JOURNAL OF MEDICAL INTERNET RESEARCH*, *24*(11). https://doi.org/10.2196/39748

Khosravi, P., & Schweitzer, M. Artificial intelligence in neuroradiology: a scoping review of some ethical challenges. *Frontiers in Radiology*, *3*, 1149461. https://doi.org/https://dx.doi.org/10.3389/fradi.2023.1149461

Li, F., Ruijs, N., Lu, Y., Li, F., Ruijs, N., & Lu, Y. (2022). Ethics & AI: A Systematic Review on Ethical Concerns and Related Strategies for Designing with AI in Healthcare. *AI*, *4*(1), 28-53. https://doi.org/10.3390/ai4010003

Maurud, S., Henni, S. H., & Moen, A. Health Equity in Clinical Research Informatics. *Yearbook of Medical Informatics*, *32*(1), 138-145. https://doi.org/https://dx.doi.org/10.1055/s-0043-1768720

McLennan, S., Kahrass, H., Wieschowski, S., Strech, D., Langhof, H., McLennan, S., Kahrass, H., Wieschowski, S., Strech, D., & Langhof, H. (2018). The spectrum of ethical issues in a Learning Health Care System: a systematic qualitative review. *INTERNATIONAL JOURNAL FOR QUALITY IN HEALTH CARE*, *30*(3), 161-168. https://doi.org/10.1093/intqhc/mzy005

Mörch, C. M., Atsu, S., Cai, W., Li, X., Madathil, S. A., Liu, X., Mai, V., Tamimi, F., Dilhac, M. A., Ducret, M., Morch, C. M., Atsu, S., Cai, W., Li, X., Madathil, S. A., Liu, X., Mai, V., Tamimi, F., Dilhac, M. A., & Ducret, M. (2021). Artificial Intelligence and Ethics in Dentistry: A Scoping Review. *Journal of Dental Research*, *100*(13), 1452-1460. https://doi.org/10.1177/00220345211013808 EA JUN 2021

Morrow, E., Zidaru, T., Ross, F., Mason, C., Patel, K. D., Ream, M., Stockley, R., Morrow, E., Zidaru, T., Ross, F., Mason, C., Patel, K. D., Ream, M., & Stockley, R. (2023). Artificial intelligence technologies and compassion in healthcare: A systematic scoping review. *Frontiers in Psychology*, *13*. https://doi.org/10.3389/fpsyg.2022.971044

Murphy, K., Di Ruggiero, E., Upshur, R., Willison, D. J., Malhotra, N., Cai, J. C., Malhotra, N., Lui, V., Gibson, J., Murphy, K., Di Ruggiero, E., Upshur, R., Willison, D. J., Malhotra, N., Cai, J. C., Malhotra, N., Lui, V., & Gibson, J. (2021). Artificial intelligence for good health: a scoping review of the ethics literature. *BMC Medical Ethics*, *22*(1). https://doi.org/10.1186/s12910-021-00577-8

Prakash, S., Balaji, J. N., Joshi, A., Surapaneni, K. M., Prakash, S., Balaji, J. N., Joshi, A., & Surapaneni, K. M. (2022). Ethical Conundrums in the Application of Artificial Intelligence (AI) in Healthcare-A Scoping Review of Reviews. *Journal of Personalized Medicine*, *12*(11). https://doi.org/10.3390/jpm12111914

Siala, H., Wang, Y. C., Siala, H., & Wang, Y. (2022). SHIFTing artificial intelligence to be responsible in healthcare: A systematic review. *Social Science & Medicine*, *296*. https://doi.org/10.1016/j.socscimed.2022.114782

Tang, L., Li, J. X., Fantus, S., Tang, L., Li, J., & Fantus, S. (2023). Medical artificial intelligence ethics: A systematic review of empirical studies. *DIGITAL HEALTH*, *9*. https://doi.org/10.1177/20552076231186064

Walk, O., Nieva, H. R., Lee, S. S. J., Elhadad, N., Walk, O. J. B. D. t., Nieva, H. R., Lee, S. S.-J., & Elhadad, N. (2022). A scoping review of ethics considerations in clinical natural language processing. *JAMIA OPEN*, *5*(2). https://doi.org/10.1093/jamiaopen/ooac039

Wang, Y., Song, Y. X., Ma, Z., Han, X. X., Wang, Y., Song, Y., Ma, Z., & Han, X. (2023). Multidisciplinary considerations of fairness in medical AI: A scoping review. *International Journal of Medical Informatics*, *178*. https://doi.org/10.1016/j.ijmedinf.2023.105175 EA AUG 2023
